# Tracking mutational semantics of SARS-CoV-2 genomes

**DOI:** 10.1101/2021.12.21.21268187

**Authors:** Rohan Singh, Sunil Nagpal, Nishal K. Pinna, Sharmila S. Mande

**Affiliations:** TCS Research, Tata Consultancy Services Ltd, Pune, India 411013; CSIR-Institute of Genomics and Integrative Biology (CSIR-IGIB), New Delhi-110025, India; Academy of Scientific and Innovative Research (AcSIR), Ghaziabad- 201002, India

## Abstract

Genomes have an inherent context dictated by the order in which the nucleotides and higher order genomic elements are arranged in the DNA/RNA. Learning this context is a daunting task, governed by the combinatorial complexity of interactions possible between ordered elements of genomes. Can natural language processing be employed on these orderly, complex and also evolving datatypes (genomic sequences) to reveal the latent patterns or context of genomic elements (e.g Mutations)? Here we present an approach to understand the mutational landscape of Covid-19 by treating the temporally changing (continuously mutating) SARS-CoV-2 genomes as documents. We demonstrate how the analogous interpretation of evolving genomes to temporal literature corpora provides an opportunity to use dynamic topic modeling (DTM) and temporal Word2Vec models to delineate mutation signatures corresponding to different Variants-of-Concerns and tracking the semantic drift of Mutations-of-Concern (MoC). We identified and studied characteristic mutations affiliated to Covid-infection severity and tracked their relationship with MoCs. Our ground work on utility of such temporal NLP models in genomics could supplement ongoing efforts in not only understanding the Covid pandemic but also provide alternative strategies in studying dynamic phenomenon in biological sciences through data science (especially NLP, AI/ML).

## 1 Background

Understanding genome sequences, an ordered collection of nucleotide bases constituting the genome, requires deciphering the rules governing the structure or positioning of the bases in the genome sequence. Human language has inherently been sequential in nature and is driven by the need for adding context (semantics) to the communication. The science of studying the rules of the human language, its grammar, semantics and more comes under the purview of linguistics^1^, and the use of Natural Language Processing (NLP) can automate this process by making computers explore, understand, learn, improve, generate, anticipate and respond to this language^2^. It does so by leveraging the fields of computer science, machine learning/ artificial intelligence and mathematics towards the common goal of understanding the language of ordered datasets. Given the structural similarities between genomic and linguistic data records, it is pertinent to ask if NLP can be utilized to decipher the hidden meanings even from ‘Genome sequence data’.

The more we explore the depths of the biological world, the greater organization we witness in the perceived complexity of biological systems^2^. It is this ‘order’ or sequential nature of various biological data that has previously inspired the use of NLP in genomics^3^, metagenomics^4^, proteomics^5^ and more. In fact, DNA, the basic hereditary unit of life, is a classic example of a sequential dataset. The use of NLP to explore the latent information of genomes is therefore a rational proposition. It becomes further interesting when the biological data is dynamic in nature, like the evolving or mutating genomes. Dynamically changing biological datasets offer the opportunity to establish a parallel with another realm of computational linguistics, namely, dynamic topic modelling^6^ and diachronic analysis of literature^7^ or language corpora. In other words, an interesting question can be posed as to whether we can treat genomes as documents (especially temporally changing documents) in order to explore and understand the continuously evolving molecular signatures (a.k.a. linguistic themes) in the vast corpus of biological features like genes or mutations (a.k.a. linguistic words).

Here, we highlight the applicability of NLP in capturing the temporal/diachronic trends in the evolution of genomic datasets. We demonstrate the same by perceiving SARS-CoV-2 genomes as documents and the associated mutations as the words of these documents. Public repositories like GISAID^8^ are teeming with SARS-CoV-2 genome sequences through sheer collaborative efforts by the scientific community. The continuation of the pandemic is only going to further add to the genomic data corpus of these public repositories. Each of these sequences is a potential variant/mutant of the original reference genome, i.e., Wuhan/WIV04/2019 (EPI_ISL_402124) requiring a deep and wide spectrum examination for understanding the viral diversity and evolution^9^. Although conventional topic modelling has been utilized to obtain insights on Covid-19 from literature data^10^, dynamic algorithms in NLP can be leveraged for understanding the diachronicity of SARS-CoV-2 mutational landscape. We demonstrate how the temporal word(mutation)-embeddings of SARS-CoV-2 nucleotide mutations can aid discovery of latent signatures (**refer Supplementary File 1**: Concepts of word embeddings, Word2Vec^11^ and DTM models, and refer **Fig 1** for graphical intuition of the same). The method enables the semantic characterization of the mutational landscape of Covid-19 and subsequent tracing of its progression with time. Such an approach could be helpful in the pathogenesis-tracking of Covid-19, especially when corroborated by leveraging the unsupervised classification of mutation vocabulary of the patient-status labelled SARS-CoV-2 genome corpora using *scattertext*^12^ (an NLP approach towards learning the mutations of concern). This methodology could also be extended to capture meaningful biological information for not only Covid-19 but other biological datasets as well. **Table 1** provides a summary of the entire approach towards the design of this research study based on asking some pertinent questions through the perspective of natural language processing.

**Fig 1:**
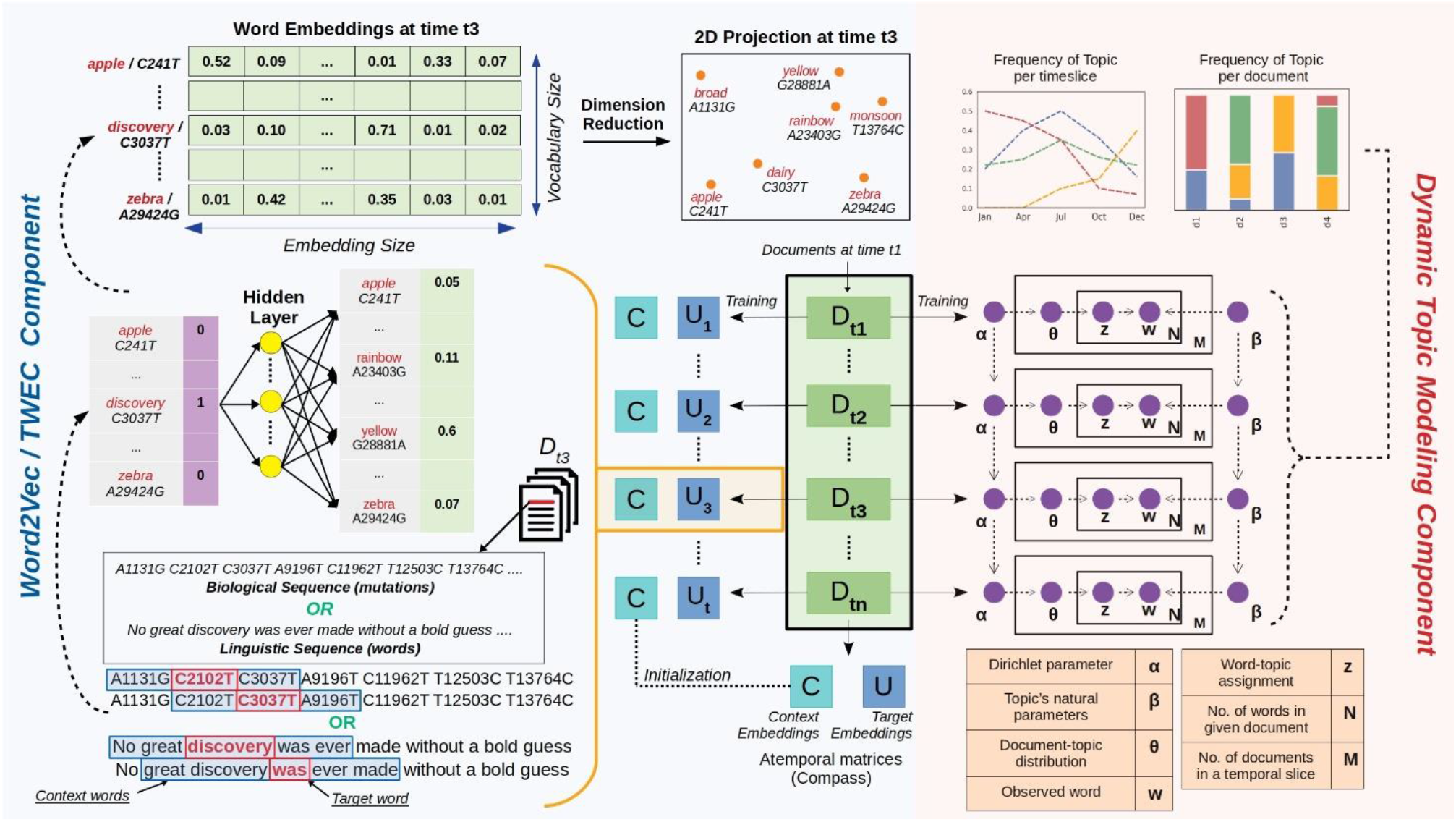
Graphical Summary of the entire NLP approach towards elucidating mutational signatures of genomic datasets (using SARS-Cov-2 genomes). Schematic representation of training procedure of both temporal natural language processing (NLP) approaches used in our study, i.e. Dynamic Topic Modeling (DTM) and Temporal Word Embeddings with a Compass (TWEC). Both techniques train on a collection of documents (corpus) that is split into respective time-slices.

**Table 1:** Summary of the questions examined towards study design for the current research

| S.No. | NLP Question | Analogous Biological Question pertaining to SARS-Cov-2 Genomes |
| --- | --- | --- |
| 1. | a) What is the vocabulary of a document?<br>b) What is the word counts in the corpus (entire documents)? | What are the mutations in a sample?<br>What is the mutation counts in GISAID dataset? |
| 2. | What are the topical themes of a document? | What signatures are present in a sample? |
| 3. | What is the distribution of topical themes in the corpus? | What is the distribution of signatures in the dataset? |
| 4. | How are the themes evolving with time? | How are the signatures evolving with time? |
| 5. | What is driving the evolution of a theme?<br>Which words are changing (semantic change) that are causing the topics to evolve? | How are signatures evolving? |
| 6. | Is there a semantic context to the word evolutions?<br>Can we trace word evolution through semantic context?<br>Can semantic drift reveal theme evolution or emergent themes? | Is mutational progression associated to genomic position? |
| 7. | How are the words changing (semantic change) w.r.t context? | How are the mutations progressing? |
| 8. | How does the semantic change of a word correspond with other words? | How does the progression of a mutation correspond with other mutations? |
| 9. | Which words can help classify documents or topics? How does word usage evolve temporally? | a) What are the characteristics mutations of Severe and Non-Severe classification?<br>b) How does the temporal clustering of these characteristic mutations look like? |

### 1.1 Establishing parallels between documents and genomics data

In order to apply NLP routines for genomic data, we need to first relate the NLP terminologies that can be utilized for obtaining insights from genome sequence dataset (**Table 2**). A genome string containing mutational information in the form of position-specific nucleotide base-pair changes corresponding to an individual/organism is akin to a ‘document’, and each mutation is analogous to a ‘word’ in a document. The entire dataset derived from GISAID^8^ can be considered equivalent to the ‘corpus’, a complete set of documents. Unique nucleotide mutation set within the entire genome datasets can be considered equivalent to the vocabulary, which refers to the entire word set present in the corpus. **Supplementary Fig1a** displays the frequency of sample (i.e., document) counts and unique mutations observed in each time-slice.

**Table 2:** Analogy between NLP terms used for documents and genomes

| S.No. | NLP Term | Definition in a Document | Analogous Genomic Relation |
| --- | --- | --- | --- |
| 1. | Word / Term | A single element of a document | A nucleotide mutation |
| 2. | Document | A sequence of words | A genome sequence containing a list of nucleotide mutations |
| 3. | Corpus | Entire collection of documents | all genomes (eg., SARS-Cov-2 genomes from GISAID dataset -- till July 2021) |
| 4. | Topic | A set of words that relate to a subject/theme. A document may comprise of one or more topics | A signature (i.e, nucleotide mutation signature) |
| 5. | Context | The particular setting or pattern in which the word occurs, usually influencing its meaning or effect | The ordered genomic position of a nucleotide mutation |
| 6. | Vocabulary | Entire collection of unique words within the corpus | Unique nucleotide mutation set within the entire dataset |

A nucleotide mutation signature is like a topic (comprised of several words) that carries a particular idea (mutations of concern). Technically, a topic is a probability distribution of its constitutive words, and as a result, documents with similar probabilities of such words to a topic signify a topic’s presence in that document. Like a document is composed of one or more topics, a genome sequence may contain more than one unique mutation signature, each having different probabilities. The inherent order in the occurrence/co-occurrence of genomic mutations is equivalent to the semantics/context of a document. A word’s meaning may differ with different texts/sentences as they are surrounded by different words (within a range of its neighbouring words called ‘window size’) and therefore influence semantics. Our study defined a ‘context’ as the genomic proximity of a mutation with other mutations, thereby establishing analogous relation between the conventional documents and genomes (**Table 2**).

## 2 Results

### 2.1 Discerning the distribution of mutational signatures (topical themes) in the SARS-Cov-2 genomes (corpus)

Six mutational signatures were inferred from the optimised model generated using Dynamic Topic Modeling (DTM) (**Fig 2**). The geographical distribution of these signatures and their mapping to various SARS-CoV-2 variants/GISAID clades are represented in **Fig 2a**. It was observed that the inferred signatures from DTM display strong coherence with ‘Variants of concern’ (VoC) and a moderate predilection towards geography. These included Signature0 that mapped to Eta and Mu variants; Signature1 mapping to the Delta variant; Signature3 (Alpha); Signature4 (Gamma, Lambda, Theta) and Signature5 (Beta). Signature2 did not map to any known VoC. The composite mutations for each of these signatures are represented in **Fig 2b**. Notably, while Signature3 (Alpha) seemed to be topically dominant in majority of the genomes collected from European nations, Signature2 showed certain prominence among Asian countries, and Signature4 was proportionally higher among South American countries. Certain countries showed single signature prominence. These included Signature2 for Japan, Signature3 for Cambodia, Bulgaria, Signature1 for Singapore, Signature0 for Saudi Arabia and Signature4 for Brazil.

**Fig 2:**
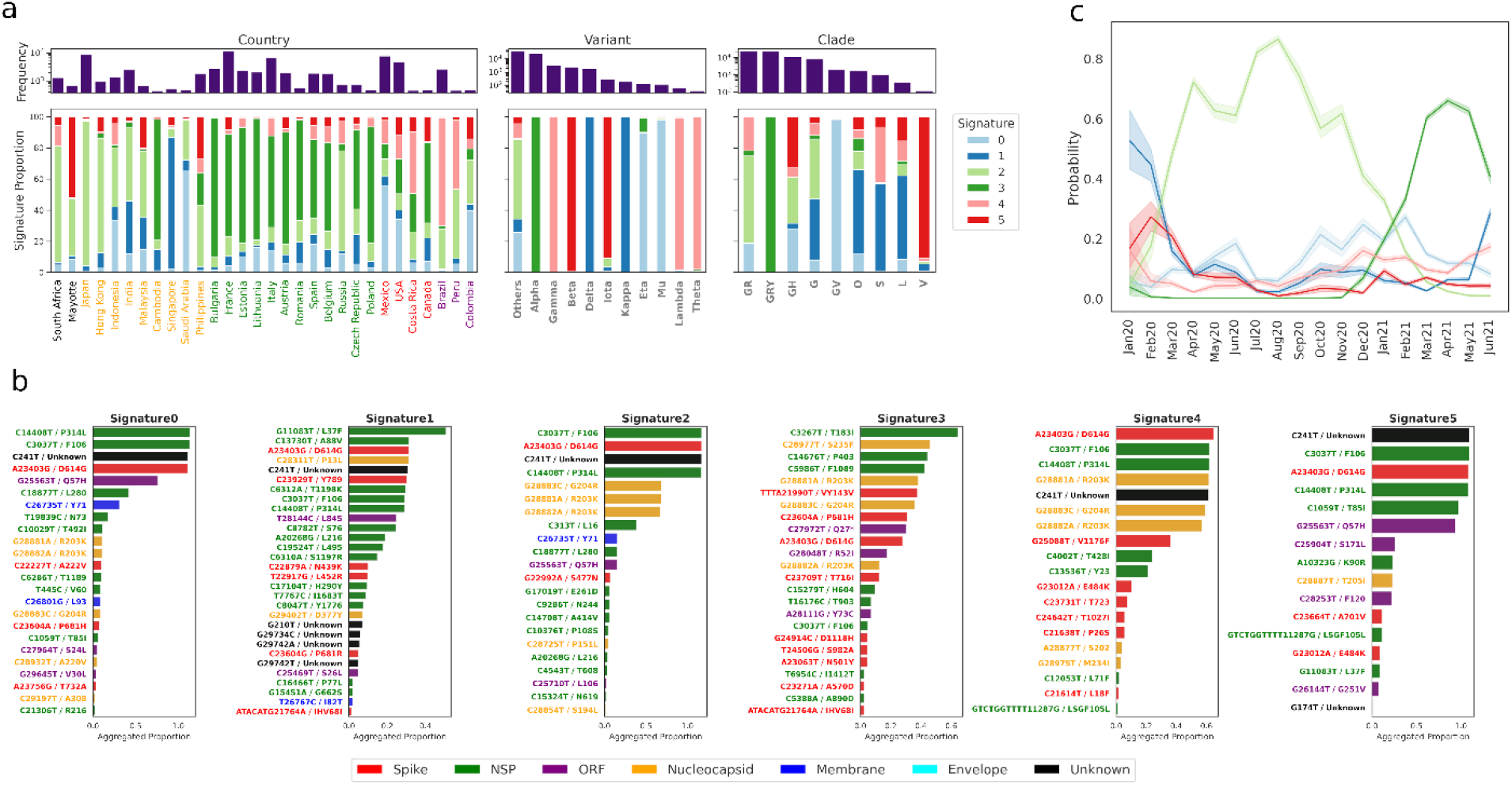
Distribution of mutational signature in SARS-Cov-2 genomes across geography. **a)** Stacked proportions of signatures (topics) in corpus segregated by 3 categories: country, SARS-Cov-2 variant and GISAID clade. The frequency of each constituent within each category are shown at the top of each stacked bar. Within the country subpanel, each country is differentially coloured according to respective continents. **b)** Aggregated proportions of word probabilities in each signature across all time-slices. Nucleotide mutations coloured in black (as Unknown) are UTRs. **c)** Temporal probabilities for each signature.

A temporal tracing of the inferred signatures is shown in **Fig 2c**. The mean probabilities of different signatures for genomes corresponding to different time-slices are plotted with a confidence interval of 0.99 (highlights the shaded area around each line). While Signature2 showed the highest probability during 2020, Signature3 (Alpha) started to manifest towards the end of 2020.

Furthermore, we probed the temporal progression of the top 20 most probable mutations within each signature (**Supplementary Fig2**). All these signatures comprised of mutations with very high initial probabilities that declined over time. Two signatures, namely, Signature3 and Signature5, had no or very few abrupt losses among their topmost probable mutations, but their corresponding mutation probabilities evened out over time. However, for the rest, new emergent words could be distinctly observed, which included Signatures 0,1 and 2, with few older mutations fading out with time. Not all signatures were found to have every word unique to itself. Certain words like those of codon G28881A, G28882A, G28883C could be seen to encompass several signatures, albeit with declining probabilities. The biological inferences/implications of DTM (and other) results are mentioned in the discussion section.

### 2.2 Collective divergence of SARS-CoV-2 mutations during different time points

To get a general overview of collective divergence of mutations during Jul2020, Dec2020 and Jun2021, 2D Uniform Manifold Approximation and Projection (UMAP)^13^ of the embedding vectors corresponding to these time points were plotted for the top 1000 most frequent mutations in the corpus. These mutations appear to converge with other mutations that share genomic proximity, i.e., those in neighbouring genomic loci or mutations belonging to the same protein (**Fig 3a)**. In Jul2020 (1st panel), the word embeddings of most frequent mutations (top 1000) were seen to be quite indistinct. This is expected as most of these mutations either had none or smaller frequencies at the beginning of 2020 and started to manifest as the pandemic spread globally (as seen in **Supplementary Fig 1a**). By Dec2020 (2nd panel), the embeddings evolved enough to gain contextual segregation which became more prominent by Jun2021 (3rd panel). For instance, from Dec2020 onwards, NSP (Non-structured Protein) related mutations were observed to cluster distinctly when their embeddings were projected.

**Fig 3:**
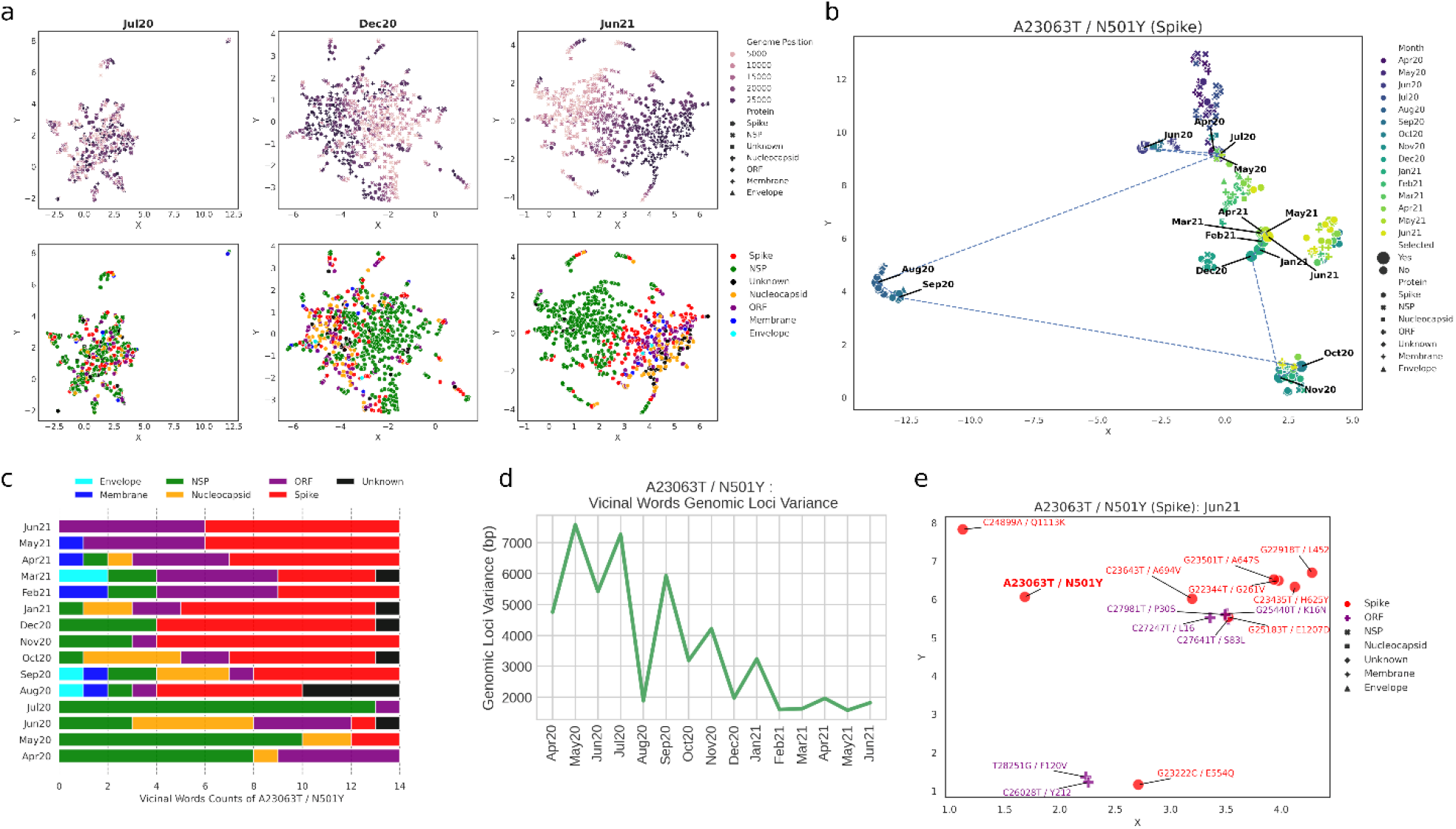
Semantic drift of mutation A23063T/N501Y. **a)** UMAP projection of word embeddings of 1000 most frequent mutations in the corpus in 3 time-slices. Most mutations tend to disperse by their genomic position. **b)** Tracking semantic drifts of mutation of concern A23063T/N501Y from Apr2020 to Jun2021, represented by the dotted line. Other points refer to the 15 most semantically related mutations in each time-slice. **c)** Protein classification of semantically closest words (neighbour) to A23063T in each time-slice. **d)** Tracking temporal shifts in the standard deviation of genomic loci of neighbouring mutations to A23063T in each time-slice. **e)** Semantically similar neighbours of A23063T in the final month of study.

### 2.3 Semantic Drift of individual Mutations of Concern (MoC)

To closely examine how the semantic of a mutation shifts temporally, four mutations of concerns (MoC) present in Spike protein, namely T22917G/L452R, A23063T/N501Y, C23604A/P681H, C23604G/P681R, were selected (as reported in outbreak.info).

The initial embedding vectors (representing meaning of a word in dynamic/continuous vector space) of A23063T/N501Y mutation for months Apr2020 to Jul2020, remained unchanged and hence closely clustered (**Fig 3b**). **Fig 3c** displays the neighbouring word composition of A23063T/N501Y for each time-slice. This mutation also displayed many non-contextual neighbours (i.e., top 15 most semantically related mutations), which were found to be mostly non-spike mutations in those initial months (as seen in **Fig 3c**). But in the months of Aug2020 and Oct2020, the embeddings changed in meaning (i.e., they experienced semantic drift), which stabilised in Dec2020 thereafter (**Fig 3b**). The vectors in the months of 2021 remained relatively unchanged with high similarity of genomic context as seen by lower genomic loci variance (*see Methods for description*), shown in **Fig 3d**, as compared to time period between Apr2020 to Jul2020, where the neighbouring words genomic loci variance were higher. Approximately half of the neighbouring mutations of Spike protein appeared Nov2020 onwards, however from Feb2021 its neighbouring mutations also constituted some from ORF protein. Given that the positional variance has been quite low (**Fig 3d**) since Jan2021, it helps to showcase that this mutation’s embedding is also linked to the ORF protein which is adjacent to Spike. **Fig 3e** shows neighbouring mutations in Jun2021.

As with ‘A23063T’ mutation, the embedding vector of C23604A/P681H mutation only gained momentum after Jul20 (**Supplementary Fig 4a)**, after which its context steadily drifted till Dec2020, post which the drift momentum as well as genomic loci variance (**Supplementary Fig 4d**) ebbed and became stabilised from Mar2021 onwards. Interestingly, its mutation neighbours from Feb2021 to Apr2021 (**Supplementary Fig 4b**) lacked those occurring in spike protein but rather had mutations from majorly Nucleocapsid and ORF protein and even envelope and membrane protein.

C23604G/P681R mutation and its counterpart mutation ‘C23604A’ shared a similar semantic transition (**Supplementary Fig 5a)**. Its semantics during initial periods remained unaltered till Jul2020, but in Aug2020, a small shift was observed, which again reverted to its previous quarter’s embedding cluster. Nov2020 and Dec2020 marked its maximum drift in semantic change, after which embeddings stabilised. The genomic loci variance dropped drastically from 2021 onwards (**Supplementary Fig 5d**), reaching the lowest at Apr2021, post which neighbouring mutations comprised majorly from ORF and Spike proteins (**Supplementary Fig 5b**).

T22917G/L452R mutation’s semantic made a sudden movement in Jun2020 but reverted close to the previous months’ embedding cluster (**Supplementary Fig 6a)**. Some drastic changes were seen in Sep2020, with neighbouring mutations being of non-spike origin, indicating non-contextual connotation (as seen by many cross markers surrounding it) (**Supplementary Fig 6b)**. Some shifts were also observed in Nov2020, wherein the neighbouring mutations were related to spike protein. After Dec2020, the embeddings stabilised in neighbourhood of mutations occurring mostly =spike protein, thereby displaying lower positional variance (**Supplementary Fig 6d)**.

In all the above-mentioned mutations, one could notice that with the progression of Covid epidemic, the neighbouring mutations comprised fewer mutations on NSP proteins, which are genomically distant to spike mutations.

### 2.4 Identifying infection severity specific mutations

NLP can also be employed on labelled datasets to classify text corpora and identify driving words/topics of such labelled documents^4^. With the aim of identifying the severity specific mutations, a total of 12,182 non-severe (‘Asymptomatic’, ‘Mild’, ‘Moderate’) infection specific (labelled) genomes and 2,497 severe (‘Critical’, ‘Severe’, ‘Fatal’) infection causing genomes were employed for the classification of latent-mutation signatures using *scattertext*. **Fig 4** displays the scatterplot of frequency of ∼2000 mutations (using *scattertext*) within ‘Severe’ and ‘NotSevere’ categories. The highlighted mutations within the figure corresponded to the top ten characteristic mutations whose likelihood of occurence was observed to be higher in one category (e.g., Severe) as compared to that in another category (e.g. NotSevere), as determined via F-scaled scores (*see methods for more details*). While all the characteristic mutations in ‘NotSevere’ category belonged mostly to NSP related mutations, the top ten characteristic mutations in ‘Severe’ category were found on several proteins. Among these, three corresponded to mutations on Spike protein (G25088T/V1176F, G22132T/R190S, A22812C/K417T), one on Nucleocapsid protein (C28512G/P80R), one on putatively non-genic mutation (G28262GAACA) and the remaining were found to be NSP linked mutations. Points in the top right corner in Fig. 4 represented the most frequent mutations (having high frequency) in both categories.

**Fig 4:**
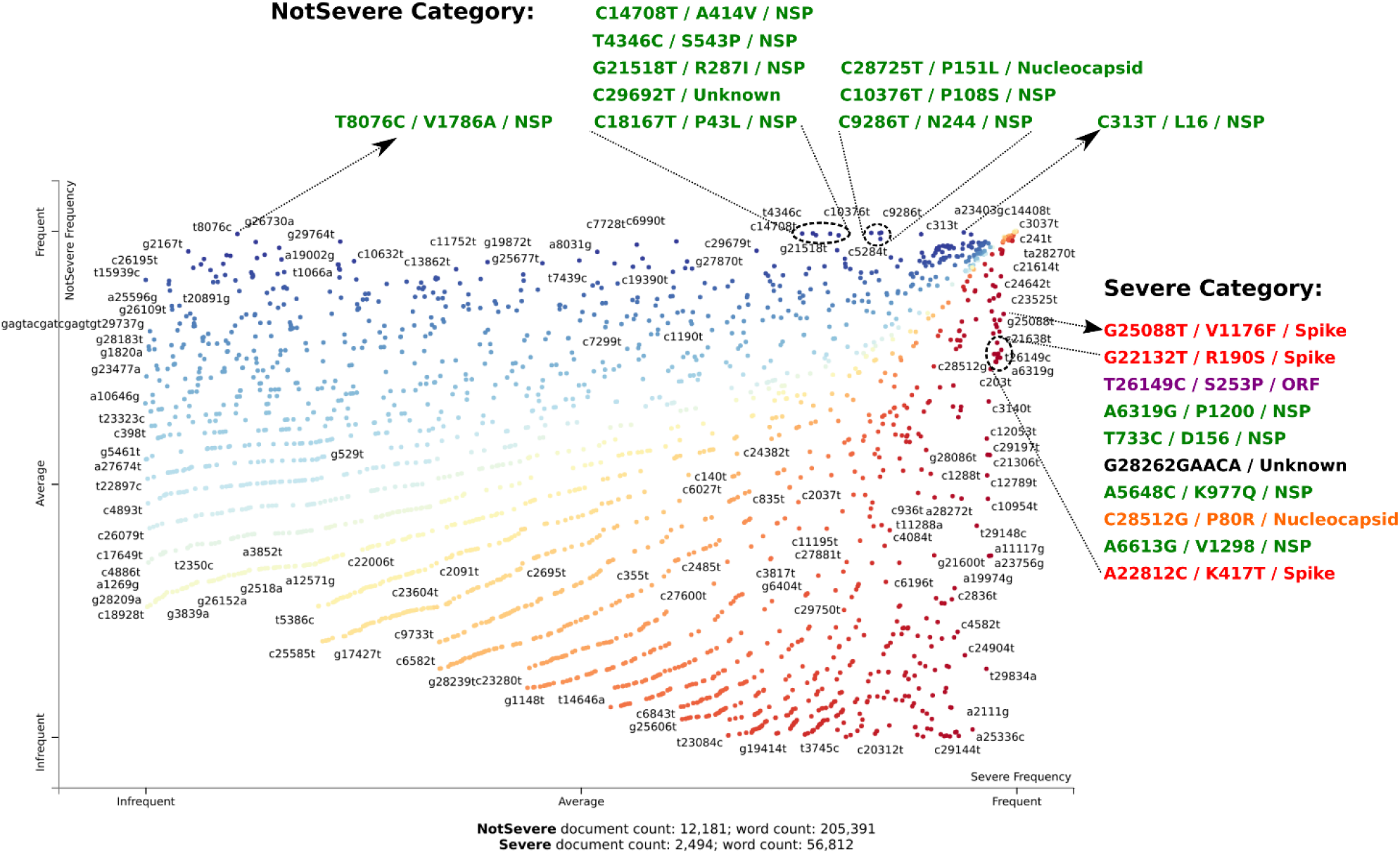
Visualising infection severity association of mutations. The figure shows the distribution of mutations in corpus along ‘Severe’ or ‘NotSevere’ frequency gradient in X and Y-axis, respectively. Points highlight mutations and are coloured according to binary classification based on F-scaled scores (see methods under Severity classification of Patient Health Status). Note that nucleotide mutations coloured in black (as Unknown) are UTRs.

### 2.5 Temporal clustering of infection severity specific mutations

To illustrate differences in temporal contextual changes in mutations associated with a physiological label (i.e., Severe or NotSevere classification), ten characteristic mutations for both categories and furthermore 30 most frequent mutations (as a neutral set) were selected for relative comparison. The mutations within the neutral set are relatively ubiquitous and non-characteristic to both infection severity categories since they are frequent in both categories, and hence they are non-markers. By semantically comparing to this neutral set, one could investigate how our derived/putative characteristic mutations converges/diverges away from a pool of mutations. K-means clustering was performed in order to observe the formation of mutation grouping in different time-slices. Cluster tracking of the aforementioned mutations (**Fig 5**) indicated varied numbers of clusters in three time-slices, namely, Dec2020, Feb2021 and May2021. Few mutations from the NotSevere category were not present in some of these time-slices, hence their embeddings were ignored for plotting for the respective time-slices. **Fig 5a-c** depict chord diagrams highlighting mutations to the respective k-means cluster (*refer Methods for further details*). **Fig 5d-f** show 2D UMAP dimension reduction plots of the embedding vectors for the three time periods of selected mutations.

**Fig 5:**
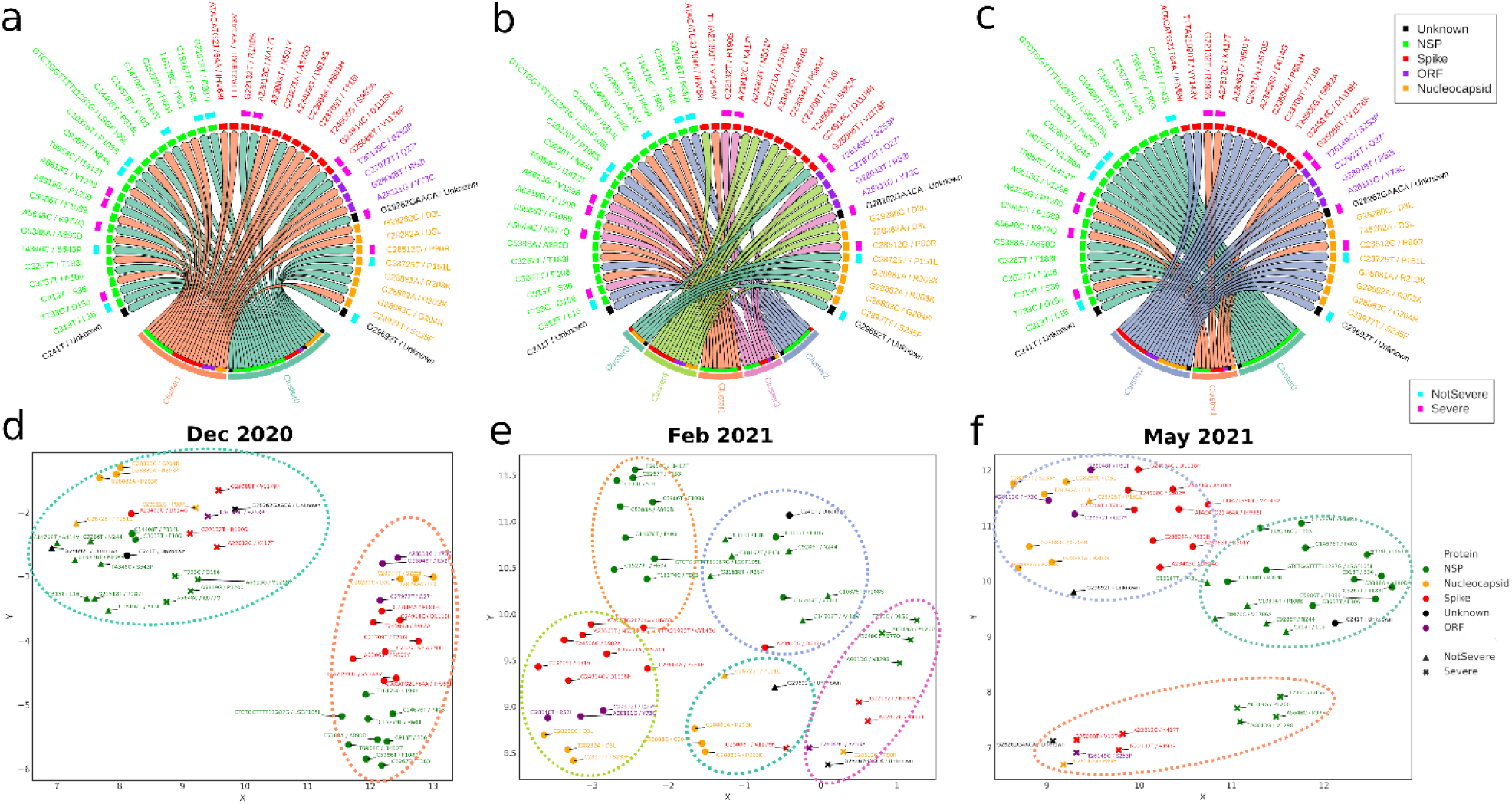
Semantic clustering of Severe, NotSevere and most frequent mutations. Panels **a,b,c** show chord diagrams of k-means clustering of 50 mutations (10 ‘Severe’, 10 ‘NotSevere’ and 30 most frequent mutations) in time-slices Dec2020, Feb2021 and May2021, respectively. Panels **d,e,f** show word embedding projection of the same mutation set in the mentioned time-slices. Mutations belonging to a cluster are encircled with dotted lines coloured according to clusters shown in panels **a,b,c**. Note that nucleotide mutations coloured in black (as Unknown) are UTRs.

In Dec2020 (**Fig 5a,d**), while the characteristic mutations of both patient health status categories (Severe and NotSevere) were noticed to be clustered into one group, the majority of the most frequent mutations were found to be clubbed into another cluster. This indicated that the semantic differences were not very prominent between mutations of both patient health status categories. In contrast, we observed unexpected segregation of mutations into numerous clusters in Feb2021, wherein k-means clustering differentiated both Severe/ NotSevere associated mutations into their respective clusters (Cluster 2, 3). However, for ‘NotSevere’ category mutations, the cluster set was not fully exclusive, indicating the specificity of signatures (the right set of co-occurring mutations) that drive a severe outcome, as opposed to those involved in an outcome which is not severe. Furthermore, we observed segregation of most frequent terms into their respective protein of origin, especially for NSP, Spike and Nucleocapsid proteins. In May2021, the trend was however found to be contrasting. Unlike the subclusters seen in the Feb2021 time-slice, fewer cluster numbers were observed. In this timepoint, the NSP mutations from NotSevere category had coalesced into a separate cluster that comprised of NSP mutations from the neutral.. Cluster 1 is solely comprised of severe-category mutations. Based on these inferences, we could suggest that the ‘Severe’ and ‘NotSevere’ mutations are semantically different as well. The biological implications of the observations have been further described in the discussion section.

### 2.6 Productivity and Frequency of Mutations of Concern (MoC) and severity associated mutations

**Fig 6** depicts the *productivity* and *frequency* for the four mutations of concern (**Fig 6a**); top 4 most frequent mutations in the corpus, namely C3037T, A23403G, G28881A and G28882A (**Fig 6b**); top 4 characteristic mutations of ‘NotSevere’ patient health category (**Fig 6c**); and top 3 characteristic spike mutations of ‘Severe’ category (**Fig 6d)**. While C3037T is a synonymous mutation providing no known evolutionary advantage for the virus, A23403G/D614G has been speculated to increase viral infectivity and reduce spike shedding^14^. G28881A and G28882A consecutive mutations have previously been speculated to affect the molecular flexibility of N protein^14^. While frequency represents the occurrence of mutations (terms) with time, productivity highlights the associability of the mutations (terms) to other mutations. The Productivity plot pattern of a term may be non-identical to its corresponding frequency plot, as a term might associate more or less to other terms irrespective of its own recurrence with time. A noticeable difference can be seen for ‘NotSevere’ mutations (**Fig 6c**), wherein their frequencies were observed to drop drastically after their peak in summer 2020. Their productivity showed retention of peaks during late 2020, post which it dropped subsequently. These therefore belonged to ‘declining terms’ wherein both frequency and productivity had reduced. Contrastingly, the frequency and productivity of ‘severe’ mutations were seen to attain a peak in 2021 itself. Frequency overlap could be observed between the top 4 most frequent mutations, which indicated how prevalent these mutations were during the pandemic and were also native to most geographies and/or lineages. Interestingly, the productivity of G28882A/R203K mutation became stagnant, which indicated that it created no new associations but solely paired with its codon partners (G28881A/R203K).

**Fig 6:**
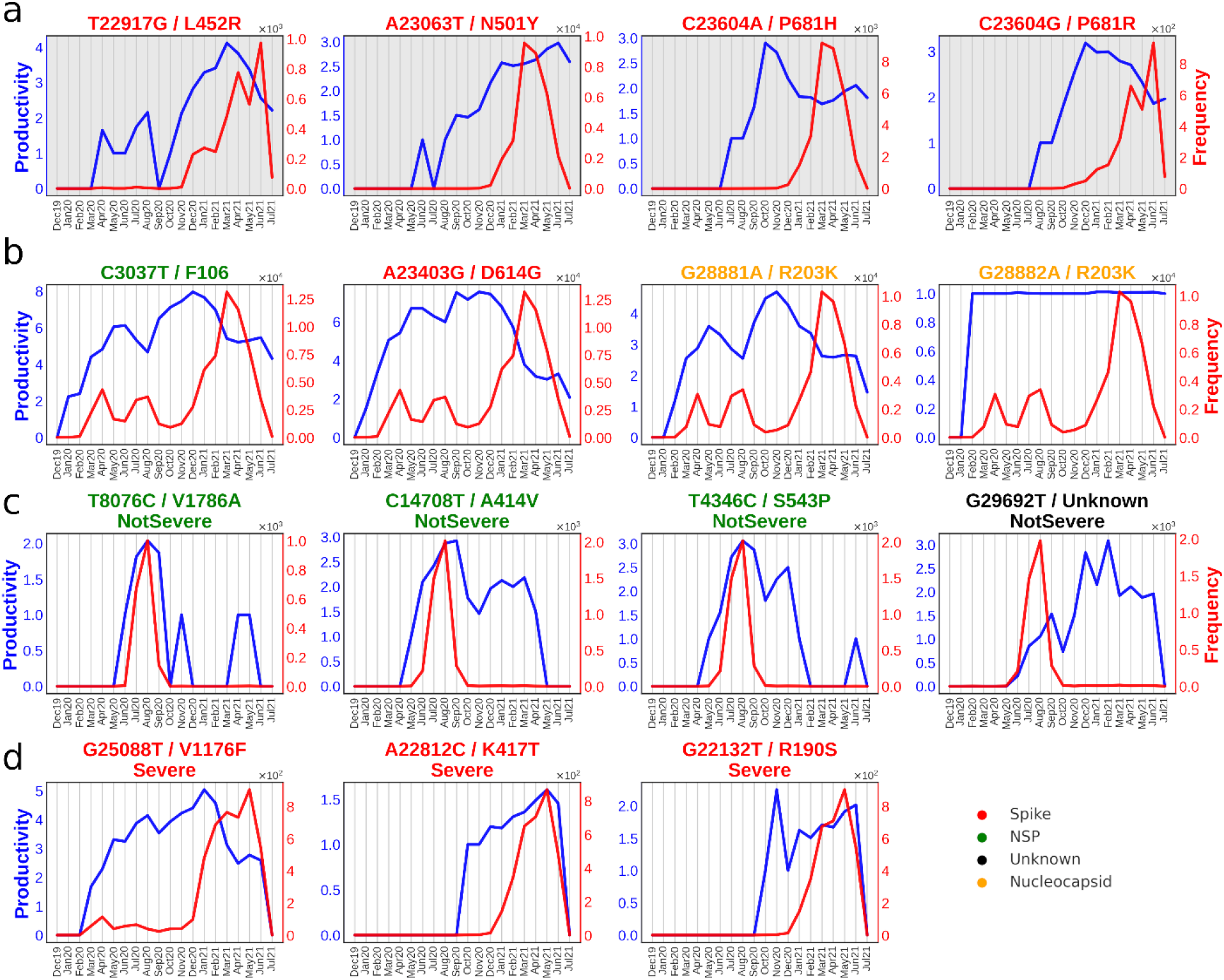
Temporal Frequency and Productivity of selected 15 mutations. Red and blue lines indicate frequency and productivity (tendency to form multi-word associations) of mutations. **a)** comprises four mutations of concern (MoC). **b)** comprises of four most frequent mutations in the corpus. **c)** and **d)** panels comprise of mutations of the ‘NotSevere’ and ‘Severe’ categories, respectively. Note that nucleotide mutations coloured in black (as Unknown) are UTRs.

Comparing the productivity and frequency of mutations of concern to their corresponding semantic drifts (**Fig 3b-e and Supplementary Fig. 4-6**) indicated that the frequency pattern didn’t relate to how the semantics of the mutations changed. However, the productivity provided following additional hints to its semantic progression.

1. *A23063T/ Spike-N501Y*: Just like in semantic drift, the productivity and frequency were relatively low/unchanged during the initial months, and the productivity steadily increased post Aug2020. The frequency had a sharp rise in Jan2021 (**Fig 6a**).
2. *C23604A/ Spike-P681H*: Just like with other mutations of concern, this mutation’s productivity started around mid-2020, after which it kept a steady rise till productivity=3 towards the end of the year, which approximately coincided with the drifting pattern displayed in semantic drift. But unlike A23063T, its productivity decreased from Nov2020 onwards to productivity=2, just when its frequency started to pick up rapidly. This peculiar observation may be due to its consistency in finding the same set of partners to which it associates. Jan2021 was found to be the month when it increased sharply for unnormalized frequency.
3. *C23604G/ Spike-P681R*: Its productivity pattern was found to be similar to its counterpart mutation, C23604A, wherein both of them showed a similar pattern of growth. However, its decline in productivity was not seen to be as sharp and sudden compared to C23604A. Also, C23604G lagged behind in frequency rise, wherein it started to make a sharp increment after Jan2021.
4. *T22917G/ Spike-L452R*: Here, the productivity pattern was observed to start much sooner than the rest of the mutations of concern. Despite low frequency during the time-slices of Mar2020 to Aug2020, it displayed a certain rise in productivity. A sudden collapse in Sep2020 occurred, after which it garnered high productivity, which steadily rose to a value of 4 in Mar2021 and then steadily declined thereafter. Its frequency curve indicated a slight rise in Dec2020, but a much higher rise was after Mar2021.

### 2.7 Acceleration of A23063T with associated mutations

Acceleration compares the similarity between two terms (mutations in our case) and determines their convergence level or divergence between two-time points. Heatmap of accelerations between the mutations obtained from DTM Signature3 (the signature that contained the MoC ‘A23063T/Spike-N501Y’ in at least one of the months) for the time period Jan2021 to Jun2021 is shown in **Fig 7a** and **Fig 7b**. To exemplify temporal changes occurring between mutations belonging to the same signature set (i.e., a DTM topic), the acceleration of signature3 mutations between two time-slices were plotted (**Fig 7a)**. Interestingly, three mutations within this signature set were semantic drift neighbours of A23063T/Spike-N501Y’ mutation (referred to as ‘SD neighbour’). These neighbours refer to mutations that were most recurrent as neighbouring mutations of A23063T from Oct2020 to Jun2021 (when productivity of A23063T started increasing rapidly).

**Fig 7:**
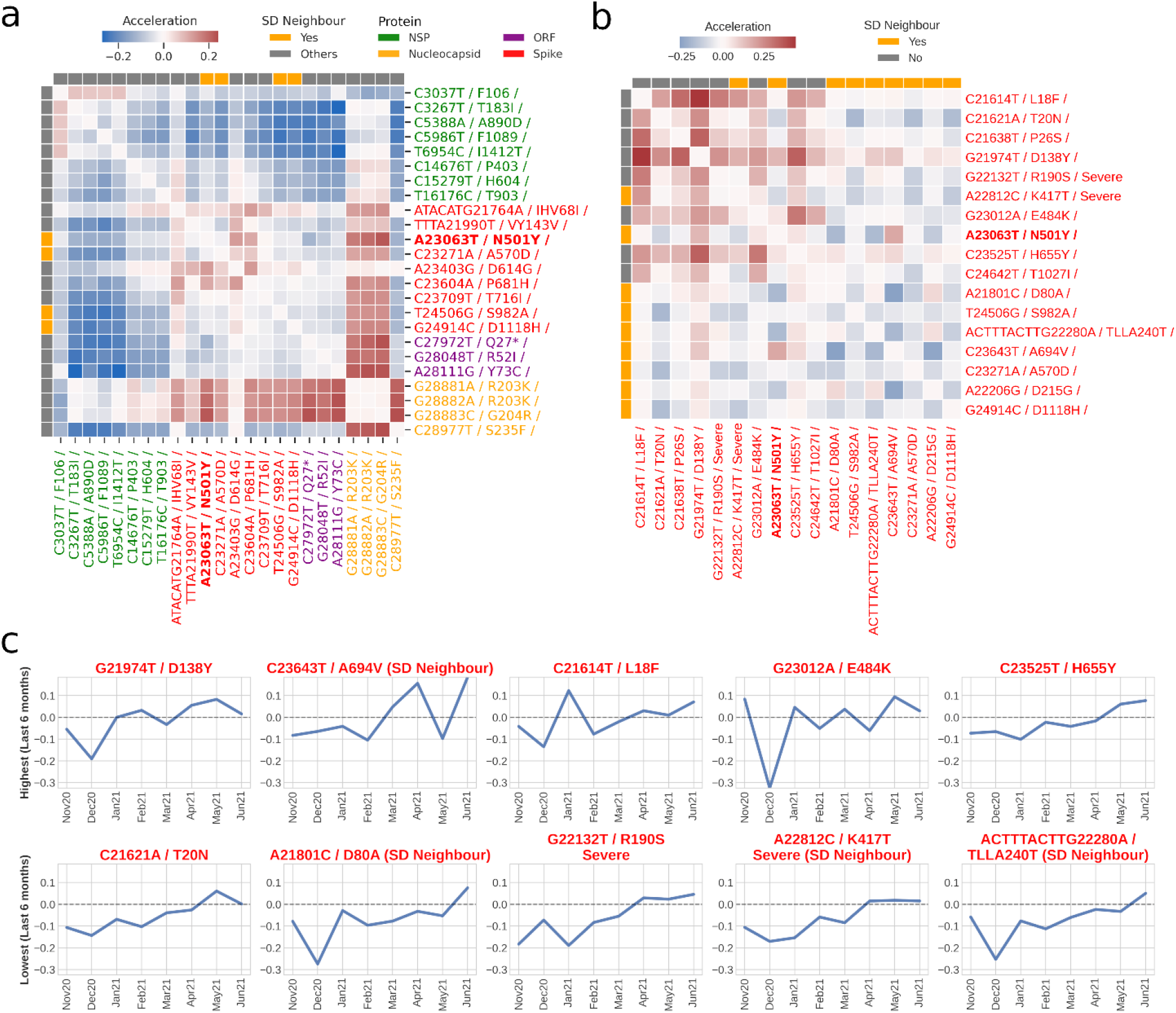
Semantic Acceleration of A23063T with associated mutations. **a)** Heatmap of semantic accelerations between Jan2021 and Jun2021 among mutations identified from signature3 of the dynamic topic model. This signature consisted of mutation of concern A23063T. ‘SD neighbour’ refers to mutations that were most recurrent as neighbouring mutations of A23063T from Oct2020 to Jun2021. **b)** Heatmap of semantic accelerations between Jan2021 and Jun2021 among key mutations of Lineage P.1^15^ and nine SD neighbours of A23063T. **c)** Dynamic acceleration tracking of ten mutations from panel (b) against A23063T. The top row highlights five mutations with the highest sum of accelerations between Jan2021 to Jun2021. The bottom row highlights five mutations with the lowest sum of acceleration between Jan2021 to Jun2021.

Broadly, while the mutations of NSP protein (coloured in green) had diverged from mutations on other proteins, the mutations of Nucleocapsid protein (coloured in yellow) had a certain level of positive acceleration with non-Nucleocapsid mutations (**Fig 7a**). A majority of the mutations on the spike protein remained largely unchanged relative to each other. It was also observed that many mutations alongside their neighbours had zero acceleration (as shown by white patches) with respect to each other. This may highlight their strong affinity to remain clustered within that duration. These mutations included the codon affecting mutations (G28881A/N-R203K, G28882A/N-R203K, G28883C/N-G204R), all of which did not diverge (i.e., zero acceleration), which also supported the observation of productivity=1 for G28882A/R203K

For demonstrating key mutations associated with one MoC, namely, A23063T/Spike-N501Y, we selected mutations of Lineage P.1^15^ (Gamma VoC) and the neighbouring mutations derived from A23063T’s semantic drift and plotted their pairwise acceleration among themselves (**Fig 7b**). Almost zero accelerations for neighbouring terms (in orange bars) derived from the semantic drift of A23063T were observed. In other words, they did not accelerate or had diverged from a semantic point of view. However, we could observe a stronger level of acceleration (seen as red cells in the heatmap) between mutations of Gamma variant (highlighted in grey). These could be prominently observed between several spike mutations, such as C21614T/L18F and G21974T/D138Y. C21614T/L18F notably has previously been reported for its impact on neutralizing some antibodies and hence contributing to the immune escape of SARS-CoV-2 mutant^16^. G21974T/D138Y lies on the N-terminal domain of S1 subunit of spike protein, however no prior report could be traced towards the evidence of its significance.

**Fig 7c** shows acceleration changes of ‘A23063T’ with the neighbouring mutations derived from A23063T’s semantic drift as well as mutations from the Lineage P.1^15^. Acceleration at each time-slice indicated a change in word embedding in comparison to the previous time-slice/month. The top row highlights mutations whose sum of acceleration over the previous six months was the highest (from Jan2021 to Jun2021). From these graphs, we could specifically identify the time-slices where the mutation pair came closer or diverged semantically. For example, C23643T/A694V had accelerated positively in March and April 2021; mutation C21614T/L18F had accelerated in Jan2021; while mutation G23012A/E484K diverged largely in Dec2020. In contrast, the mutations with the lowest sum of accelerations (between Jan2021 to Jun2021) with MoC ‘A23063T’, were found to have slightly diverging trend initially, but with time the trend approached zero acceleration (i.e., unchanging semantics).

## 3 Discussion and Conclusion

Dynamic topic modeling (DTM) approach deployed in our study can aid evolution informed viral classification. It can assist in the identification of mutations and their temporal prominence as key drivers of epidemiological or biological events when associated with a parameter of a labelled dataset (such as disease severity and geography) (**Fig 2**).

Previously^10^, we used (static) topic modeling to identify signature mutations for different geographies (**Supplementary Fig 7,8**). But while static topic modeling is a resourceful unsupervised method for identifying signatures, it does not however provide diachronic insights into the frequency of any signature. Hence, DTM provides additional depth by showcasing how a variant evolves and what mutations drive this evolution (**Fig 2**).

The signatures may not have an apparent phylogenetic relationship but can be influenced by linguistic/semantic. In other words, the collective corpus directs the classification rather than genomic variants branching from phylogeny. Thus, the DTM methodology is a biology agnostic model which is capable of finding biological signatures. For instance, the Alpha variant, which originated in the United Kingdom sometime in Sep2020, became an epidemiological concern with rising cases across Europe by late 2020^17,18^. The same has been captured quite effectively by the DTM model (**Fig 2a** and **2b**). Other geography-specific observations can be seen for the Gamma variant that originated in Brazil and has a very high proportion in variant distribution of Brazil. Similarly, the Delta variant, which originated in India (having smaller but with discernible proportion) but having significant presence in Singapore, which was one of the first countries outside India with detected delta variant. Capturing such trends without prior clinical information hints towards the suitability of these NLP-related approaches in assisting the discovery/tracing of VoCs and MoCs.

At an individual level, mutations commonality across signature/variant can also be tracked and ascertained via linguistic approaches. In our study, deploying TWEC model offered a neoteric approach to see not just the mutation-mutation relationship but also their evolution (**Fig 3, 5, 7**). To showcase its applicability, we focussed on semantic changes in mutations of concern (MoC) by considering them as words in a document. One prominent mutation of concern, A23063T/Spike-N501Y, has been purported to increase transmission by variants that harbour this mutation^19^. It is predominantly higher in Alpha and Beta variants (shown in **Supplementary Fig 1b**), both of which became major concern around Dec2020^19,20^. **Fig 6** shows the frequency and productivity of a few key mutations analysed in our study. Here we can observe that frequency of A23063T is detectable very strongly from Dec2020 onwards. However, the increment in productivity predates that of frequency, indicating that it prevailed prior to becoming a part of concerning variants, where its productivity peak emerged from Aug2020 onwards. We can also observe in **Fig 3c** that before Aug2020, the neighbouring mutations were mostly of non-Spike origin, possibly owing to initialised random word vectors due to smaller mutation vocabulary in early time-slices.

Another aspect in our study pertains to utilising labelled patient status annotation of a document’s (i.e., genome sample) affiliation to a binary category, ‘Severe’ and ‘Non-Severe’. With the help of F-scaled scores computed using the *scattertext* package^12^ (*refer methods for further details*), we were able to classify certain mutations that correlate to these two classes. We displayed the top 10 mutations for each class in **Fig 4**. Examples of delineated ‘Severe’ mutation are G25088T (corresponding to V1176F of Spike protein) and T26149C (corresponding to S253P of ORF3 protein), which have also been previously reported as mortality linked mutations21s21. Two other Spike mutations of the ‘Severe’ category, G22132T (R190S) and A22812C (K417T) are shown to be key members of Lineage P.1^15^. Lysine417 role in interaction with ACE2 receptor for infectivity^15,22^ obliquely substantiates our severity association, although further investigation would be needed to justify this linkage. Hence, this category-linked linguistic method can provide useful perspectives related to that class, which in our case related to disease severity and mutations, which can be corroborated further from a structural point of view.

In the present study, we showcase NLP-driven yet biologically unconventional methods for trailing the evolutions of variants of concern and the associated mutations of concern. We believe that using genomic datasets from a linguistic perspective could supplement the ongoing efforts in understanding associations between the mutations of SARS-CoV-2 genomes. The different NLP-methodologies undertaken, namely DTM and TWEC, can provide temporal insights from a regional or global perspective that may assist in variant identification, classification or even bring forth structural connections for therapeutic targeting.

## 4 Methods

### 4.1 Data Preparation

SARS-CoV-2 metadata was downloaded from GISAID, which at the moment of collection (Jul2021) contained ∼2.6 million samples. All the samples without patient health status were filtered out. Among these 77,283 filtered samples, samples whose collection date was complete were selected. This led to a total of 75,084 samples comprised of ∼30,000 mutations.

To profile those mutations, we used a previously adopted protocol23 adopted protocol23 adopted protocol23. Here, fasta files of individual genomes samples were downloaded and mapped with NCBI GenBank accession NC_045512 (GISAID ID EPI_ISL_402125 / coronavirus-2 isolate Wuhan-Hu-1) as the reference (Reference.fa) using minimap2^24^ (with default parameters other than ‘--cs -cx asm5‵). Subsequently, paftools.js was used for identifying nucleotide variations (.vcf file) with respect to reference. Amino acid changes corresponding to the identified nucleotide variations were predicted using BCFtools/csq program^25^. All genome samples were binned into different time-slices, where each time-slice corresponded to months from Dec2019 to Jul2021, bringing to a total of 18 time-slices (as shown in **Supplementary Fig 1a**).

### 4.2 DTM training

DTM models were created with different ‘k’ values (no. of topics) from 4 to 15 while keeping the rest of the parameters to default values. Unlike classical LDA, wherein the topic remains unchanged and thus choosing optimal ‘k’ value from coherence score is rather straightforward. Therefore, for DTM implementation, the ‘k’ value was chosen based on how distinct each topic was in each timeframe. For that, a similarity metric was created that measured the difference in the top 20 most probable mutations between different topics at a given timeframe. This was again measured using the Jaccard similarity metric, which was employed in hyperparameter training for the Skip-gram model. For a particular value of ‘k’, the Jaccard similarity between any two topics is summed across all timeframes and divided by the number of time-slices. Then this Paired Jaccard score is summed for all pairs of topics possible for the value of ‘k’ and divided by the total no. of pairs.

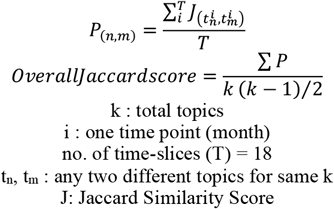

From **Supplementary Fig 3a**, k=6 was chosen as the topic number since its inter-topic similarity was low (with the sharpest drop) as well as fewer topics make it more interpretable for tracking in drift analysis.

### 4.3 DTM Parameters

For DTM, each sample from the GISAID dataset till the month of July 2021 was considered as a document (See Data preparation for further details). Nucleotide mutation lists from GISAID samples were segregated according to their month of collection. These constituted the time-slices on which the DTM was performed with ‘k’ (no. of topics) = 6, lda_sequence_min_iter=6, alpha=0.01, rng_seed=0. A python wrapper for Dynamic Topic Models (DTM) present in gensim^26^ python package (version 3.8) was used on top of compiled binaries of DTM from the original paper^6^ to execute DTM on our corpus. Signature distribution (**Fig 2a**) were subsequently calculated by computing the signature distribution (i.e., probabilities of individual signature) for each document (i.e., genome sample) and grouping them into respective countries/variants and then calculating the overall proportion of signature probabilities for each. Aggregated probabilities of ‘most probable mutations’ in each signature (is shown in **Fig 2b)** were computed by summing probabilities in all time-slices, and only the top 20 mutations with the highest sum of probabilities are listed.

### 4.4 DRIFT Skip-gram Training

To build our temporal TWEC model and perform subsequent analysis, we modified and utilized the DRIFT toolkit^7^. Hyperparameters of the Skip-gram architecture of a Word2Vec model like embedding size (i.e., vector dimensionality), word size (i.e., context window) and number of negative samplings affects the quality of the model. In order to narrow down towards an optimal set of hyperparameters, a similar approach that was incorporated in Dridi et al.^28^ was followed. The approach is to find overlap in the closest words to a target word with varying hyperparameters, yielding models that were trained on different training sets. The overlap (between models trained on two training sets) is measured via Jaccard similarity between the closest neighbouring words to a target word from each timeframe obtained from one training set vs the other. In our case, the top 5 nearest neighbours were chosen for stringent similarity testing, and the top 50 most frequent words were chosen as the set of target words for whom the Jaccard similarity metric was computed.

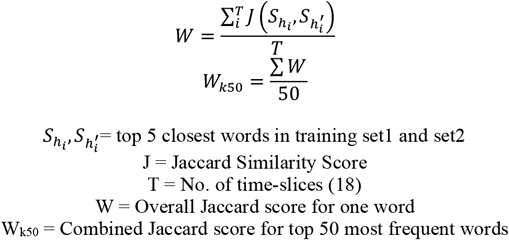

On this basis, the training parameters were obtained, specifically for Embedding Size, Window Size, and Negative Sampling. Each pair of training sets whose overlap is measured via the ‘Overall Jaccard score’ for a word differed only by one hyperparameter value. The training ranges are shown in **Supplementary Tables 1,2,3**; wherein training parameters ranged from 50-400 for Embedding Size, 2-60 for Window Size, and 1-21 for Negative Sampling. The ‘Overall Jaccard score’ for top 50 most frequent words (W_k50_) can be seen in **Supplementary Fig 3b, 3c, 3d** for Embedding Size, Window Size, and Negative Sampling, respectively.

For embedding Size, the window size of 10 was first randomly fixed and different training parameters were utilized for embedding size. For embedding size, the mean value of Overall Jaccard score (W_k50_) increases rather sluggishly after embedding size of 100. The embedding size was fixed at a value of 200 so as to not make the training too computationally intensive and time-consuming. Similarly, word size of 8 was chosen, as no significant change can be observed beyond that, and a smaller word size helps preserve the genomic context. A negative sampling of 14 was chosen as it showed a relative maximum to other values. Therefore, the final model selection had the parameters as Word Size=8, Negative Sampling=14, Embedding Size=200.

### 4.5 Individual Semantic Drift

In this study, four mutations of concerns as reported by outbreak.info were considered. These were T22917G, A23063T, C23604A, C23604G; all belonging to Spike protein. UMAP^13^ was used for dimension reduction of individual mutation’s embedding vectors for each time-slice. Additionally, 15 closest similar mutations (as determined through cosine similarity) were chosen for each of those time-slices and were plotted as well. Distribution of neighbours (horizontal barplot) for each MoC was made by mapping which protein the neighbouring mutations (i.e., 15 closest semantically similar mutations) for each time-slice originate from and plotting them in **Fig 3c** and **Supplementary Fig 4,5,6**.

### 4.6 Genomic loci variance

Genomic loci variance showcases the deviation in genomic positional distance to the MoC for each time-slice. It is calculated by first extracting the genomic loci position of each neighbouring mutation in a time-slice and computing the standard deviation of the absolute differences between the genomic position of MoC with its neighbouring mutations.

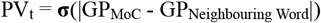

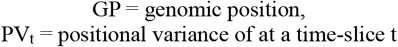

### 4.7 Productivity

To understand the diachronic development of a term/word, Schuman et al.^27^ described a method to model the life cycle of individual words with the help of a term’s frequency and productivity. ‘Term frequency’ signifies the frequency of occurrence of a given term in a given time-slice. In contrast, ‘Term productivity’ measures the ability of a single term to produce new, related multi-word terms. They have conceptualized term productivity by calculating the entropy of conditional probabilities of all *n* multi-word terms *m* that contains single-word term *t* for each timepoint *y*. This is represented in the equation below:

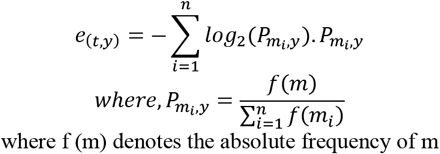

Collectively they yield three types of term dynamics:

1. Growing terms exhibit an ongoing increase of both productivity and frequency
2. Consolidated terms still grow in frequency, but not in productivity
3. Terms in decline seem to have reached an upper bound of productivity and are being used less in terms of frequency.

### 4.8 Acceleration

To highlight the nature of similarity and the extent of convergence/divergence between two words across two time periods, the word-pair ‘acceleration’ was used. By finding the difference in the cosine similarity of the embedding vectors for a pair of words between two timeframes, the acceleration with which the corresponding word-pair was getting closer (or not) were computed. In other words, it was examined whether they are appearing more frequently in similar contexts or separating out of that context. This metric has been formalised by Dridi et al.^28^ and is described below.

All distances between two words word_i_ (w_i_) and word_j_ (w_j_) are calculated by the cosine similarity between embedding vectors 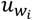 and 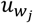

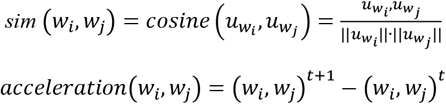

Dynamic acceleration plot **Fig 7c** shows changes in acceleration between two words at every time-slice. In the present case, dynamic acceleration was calculated only for MoC with their semantic drift neighbours. However, only ten neighbouring words/mutations were selected to display in the graphs, five of them being the most positively accelerating mutations and the other five being the lowest accelerating mutations. The most accelerating neighbour wordset and lowest accelerating wordset were chosen by calculating the sum of accelerations in the past six months (i.e., Jan2021 to Jun2021) and choosing the top five and bottom five, respectively.

### 4.9 Severity classification of Patient Health Status

Scattertext package^12^ version 0.1.4 was used in python3 environment. It uses a criterion called ‘Scaled F-Score’ to find how associated words are with categories. Terms associated to a category must have high category-specific precision and category specific frequency (percentage of terms in the category that are term). F-score is thus the scaled harmonic mean of this precision and frequency.

Scaled F-score ranges from -1 to 1, wherein words closer to -1 (red-coloured) or 1 (blue coloured) are more characteristic of category1 or category2, respectively. On the other hand, a word is plotted according to its frequency in both categories, i.e., its cartesian coordinates are its frequency (per 25,000) in category1 (i.e., the X-axis) and frequency (per 25,000) in category2 (i.e., the Y-axis). Points corresponding to terms are selectively labelled so that they don’t overlap with other labels or points. The minimum words frequency for plotting was set to 5, resulting in 2271 words.

### 4.10 Tracking Clusters

Many words collectively form a context, i.e., they are similar in meaning. But as each of the words drift, so do the semantic cluster they were part of. Therefore, one can track the transition of several words (i.e., mutations) as to whether they consolidate in their semantic context or disunite with time. Here for each time-slice, mutations were clustered by first reducing the dimensions of their word embeddings using UMAP dimension reduction, then clustering them using faiss’s library for k-means clustering algorithm. The optimal number of clusters for each time-slice was determined using *Silhouette Score* module from the sklearn package^29^. The resulting k-means cluster demarcation for a set of chosen mutations for a given month is represented using circlise package in R language. Furthermore, the UMAP projection of word embeddings of those mutations for a given month was also plotted.

## Data Availability

All data produced in the present work are contained in the manuscript. Genome sequences and metadata employed in this research are available on gisaid.org

https://www.gisaid.org

## Acknowledgement

We gratefully acknowledge all the Authors from the Originating laboratories responsible for obtaining the specimens and the Submitting laboratories where genetic sequence data were generated and shared via the GISAID Initiative, on which this research is based. Genome sequences and meta-data should be downloaded from https://www.gisaid.org. We gratefully acknowledge the original contributors of the virus genome sequences used in this study in Supplementary File 2.

## Author contribution

SN conceived the idea. RS, SN designed the study. NKP, SN gathered the data. NKP did the variant calling. RS performed model construction, training and optimization. RS, SN performed the analyses. RS wrote first draft of manuscript and designed figures. SSM, SN supervised the work and finalized the manuscript. All authors reviewed and approved the submission.

## REFERENCES

1. Hammarström, H. Linguistic diversity and language evolution. Journal of Language Evolution 1, (2016).

2. Brooks, D. R., Collier, J., Maurer, B. A., Smith, J. D. H. & Wiley, E. O. Entropy and information in evolving biological systems. Biology and Philosophy 4, (1989).

3. Yandell, M. D. & Majoros, W. H. Genomics and natural language processing. Nature Reviews Genetics vol. 3 (2002).

4. Liu, L., Tang, L., Dong, W., Yao, S. & Zhou, W. An overview of topic modeling and its current applications in bioinformatics. SpringerPlus 5, 1608 (2016).

5. Ofer, D., Brandes, N. & Linial, M. The language of proteins: NLP, machine learning & protein sequences. Computational and Structural Biotechnology Journal vol. 19 (2021).

6. Blei, D. M. & Lafferty, J. D. Dynamic topic models. in ACM International Conference Proceeding Series vol. 148 (2006).

7. Sharma, A., Chhablani, G., Pandey, H. & Patil, R. DRIFT: A Toolkit for Diachronic Analysis of Scientific Literature. arXiv preprint 2107.01198 (2021).

8. Shu, Y. & McCauley, J. GISAID: Global initiative on sharing all influenza data – from vision to reality. Eurosurveillance vol. 22 (2017).

9. Rambaut, A. et al. A dynamic nomenclature proposal for SARS-CoV-2 lineages to assist genomic epidemiology. Nature Microbiology 5, (2020).

10. Nagpal, S., Srivastava, D. & Mande, S. S. What if we perceive SARS-CoV-2 genomes as documents? Topic modelling using Latent Dirichlet Allocation to identify mutation signatures and classify SARS-CoV-2 genomes. bioRxiv 2020.08.20.258772 (2020) doi:10.1101/2020.08.20.258772.

11. Mikolov, T., Chen, K., Corrado, G. & Dean, J. Efficient estimation of word representations in vector space. in 1st International Conference on Learning Representations, ICLR 2013 - Workshop Track Proceedings (2013).

12. Kessler, J. S. ScatterText: A browser-based tool for visualizing how corpora differ. in ACL 2017 - 55th Annual Meeting of the Association for Computational Linguistics, Proceedings of System Demonstrations (2017). doi:10.18653/v1/P17-4015.

13. McInnes, L., Healy, J., Saul, N. & Großberger, L. UMAP: Uniform Manifold Approximation and Projection. Journal of Open Source Software 3, (2018).

14. Justo Arevalo, S. et al. Global Geographic and Temporal Analysis of SARS-CoV-2 Haplotypes Normalized by COVID-19 Cases During the Pandemic. Frontiers in Microbiology 12, (2021).

15. Harvey, W. T. et al. SARS-CoV-2 variants, spike mutations and immune escape. Nature Reviews Microbiology vol. 19 (2021).

16. McCallum, M. et al. N-terminal domain antigenic mapping reveals a site of vulnerability for SARS-CoV-2. Cell 184, (2021).

17. Hodcroft, E. B. et al. Spread of a SARS-CoV-2 variant through Europe in the summer of 2020. Nature 595, (2021).

18. Stadtmüller, M. et al. Emergence and spread of a sub-lineage of SARS-CoV-2 Alpha variant B.1.1.7 in Europe, and with further evolution of spike mutation accumulations shared with the Beta and Gamma variants. medRxiv 2021.11.01.21265749 (2021) doi:10.1101/2021.11.01.21265749.

19. Huang, H., Zhu, Y., Niu, Z., Zhou, L. & Sun, Q. SARS-CoV-2 N501Y variants of concern and their potential transmission by mouse. Cell Death and Differentiation vol. 28 (2021).

20. Walensky, R. P., Walke, H. T. & Fauci, A. S. SARS-CoV-2 Variants of Concern in the United States-Challenges and Opportunities. JAMA - Journal of the American Medical Association vol. 325 (2021).

21. Fang, S. et al. Updated SARS-CoV-2 single nucleotide variants and mortality association. Journal of Medical Virology 93, (2021).

22. Lan, J. et al. Structure of the SARS-CoV-2 spike receptor-binding domain bound to the ACE2 receptor. Nature 581, (2020).

23. Dimonaco, N. J., Salavati, M. & Shih, B. B. Computational analysis of sars-cov-2 and sars-like coronavirus diversity in human, bat and pangolin populations. Viruses 13, (2021).

24. Li, H. Minimap2: Pairwise alignment for nucleotide sequences. Bioinformatics 34, (2018).

25. Danecek, P. & McCarthy, S. A. BCFtools/csq: Haplotype-aware variant consequences. Bioinformatics 33, (2017).

26. Reh urek, R. & Sojka, P. Software Framework for Topic Modelling with Large Corpora. in Proceedings of the LREC 2010 Workshop on New Challenges for NLP Frameworks 45–50 (ELRA, 2010).

27. Schumann, A.-K. Brave New World: Uncovering Topical Dynamics in the ACL Anthology Reference Corpus Using Term Life Cycle Information. in (2016). doi:10.18653/v1/w16-2101.

28. Dridi, A., Gaber, M. M., Azad, R. M. A. & Bhogal, J. DeepHist: Towards a Deep Learning-based Computational History of Trends in the NIPS. in Proceedings of the International Joint Conference on Neural Networks vols. 2019-July (2019).

29. Pedregosa, F. et al. Scikit-Learn: Machine Learning in Python. J. Mach. Learn. Res. 12, 2825–2830 (2011).

