## Supplementary File 1 for "Tracking mutational semantics of SARS-CoV-2 genomes"

### Identifying mutation signatures (themes) from genomic documents

In NLP, topic modeling is a type of statistical modeling approach used to describe the process of finding abstract ‘topics’ in a corpus. Latent Dirichlet Allocation[1] is one popular method that learns a predefined number of topics from a corpus. Each document can be considered a probability distribution of one or more topics, and each topic is a representation of probability distribution over certain n-grams (words) associated with them. LDA has frequently been used in biological contexts for data exploration via clustering, classification or features extraction in an unsupervised fashion[2–4].

Like LDA, Dynamic Topic Models[5] (DTM) are stochastic models but can help in analysing the evolution of ’latent’ topics within a corpus over time. In a temporal dataset, the context is heavily dependent on the order of the data (i.e., the documents) that are grouped by time-slices (e.g., days, months, years). However, a (static) LDA model does not consider this ordering. In a DTM model, the words are considered exchangeable and so is the order of appearance of documents, where it is assumed that the documents of each time-slice are generated from the topics that were evolved from the previous time-slice.

DTM associates words to topics based on their statistical relationship of occurrences, then accumulatively assigning topic probabilities to each document. It assigns what topics comprise a document. *Given a biological problem such as tracking mutations of a species, wherein a genome of an organism (i.e., a document) is evolving with time, a use case of DTM could be to discern a set of mutational signatures across a species and investigate how they change in time, essentially, does a mutation differ in its importance as a signature with time, or do we see novel signatures with time.*

### Semantic (context) Drift and Topic (signature) evolution

From a linguistic perspective, words may ‘drift’ from their semantics over time due to changing linguistic, social and cultural norms. Analogous to this, the mutational prevalence may also shift with respect to time, leading to new variants or subtyping of the disease or depicting a change in pathogenesis.

The solution to the problem of tracing the semantic change in an NLP setting is rooted in Word2Vec[6]. It works on the philosophy of deriving relations between words through their word vectors/embeddings, that forms a context to which words with similar vectors can be associated and thus, it becomes useful in either classifying innominate words or discovering contexts.

Temporal Word Embeddings with a Compass[7] (TWEC), yields similar type of inferences but with a temporal level of detail. As new documents are generated for newer timepoint, we can trace shifts in the meaning of words or associations with other words via this approach. Therefore, taking the same biological problem (as mentioned in the above section), we can track changes in genomic context of mutation occurrence.

TWEC can be modeled around two Word2vec architectures: Continuous bag-of-words (CBOW) and Skip-gram. Skip-gram predicts the context word for a given target word. The target word (mutation) vectors/embeddings can be provided as input, which can then be used to predict context words occurring nearby to it within a fixed window size (**Fig 1**). Conversely, CBOW method can take the context (as vectors/embeddings) as input to predict the word (i.e. target) corresponding to the said context. While training a TWEC model, one of the two embeddings is fixed while the other is updated with each time-slice of the corpus. The fixed embedding acts as a compass to guide the training of the temporal embeddings to their respective time-slices. The semantic shift can inherently be computed from the TWEC embeddings for a word in each time-slice. By calculating the euclidean/cosine distances for the embeddings of words in two timeframes, we can get an estimate of a word's context over time.

| Set name | Negative Samples | Window Size | Embedding size |
|---|---|---|---|
| E50 | 10 | 10 | 50 |
| E100 | 10 | 10 | 100 |
| E150 | 10 | 10 | 150 |
| E200 | 10 | 10 | 200 |
| E250 | 10 | 10 | 250 |
| E300 | 10 | 10 | 300 |
| E400 | 10 | 10 | 400 |

**Supplementary Table 1:** TWEC Training parameter for embedding-size tuning

| Set name | Negative Samples | Window Size | Embedding size |
|---|---|---|---|
| W2 | 10 | 2 | 200 |
| W3 | 10 | 3 | 200 |
| W4 | 10 | 4 | 200 |
| W5 | 10 | 5 | 200 |
| W6 | 10 | 6 | 200 |
| W7 | 10 | 7 | 200 |
| W8 | 10 | 8 | 200 |
| W9 | 10 | 9 | 200 |
| W10 | 10 | 10 | 200 |
| W11 | 10 | 11 | 200 |
| W12 | 10 | 12 | 200 |
| W13 | 10 | 13 | 200 |
| W14 | 10 | 14 | 200 |
| W15 | 10 | 15 | 200 |
| W17 | 10 | 17 | 200 |
| W20 | 10 | 20 | 200 |
| W23 | 10 | 23 | 200 |
| W25 | 10 | 25 | 200 |
| W27 | 10 | 27 | 200 |
| W30 | 10 | 30 | 200 |
| W35 | 10 | 35 | 200 |
| W40 | 10 | 40 | 200 |
| W45 | 10 | 45 | 200 |
| W50 | 10 | 50 | 200 |
| W60 | 10 | 60 | 200 |

**Supplementary Table 2:** TWEC Training parameter for word-size tuning

| Set name | Negative Samples | Window Size | Embedding size |
|---|---|---|---|
| NS1 | 1 | 8 | 200 |
| NS2 | 2 | 8 | 200 |
| NS3 | 3 | 8 | 200 |
| NS4 | 4 | 8 | 200 |
| NS5 | 5 | 8 | 200 |
| NS6 | 6 | 8 | 200 |
| NS7 | 7 | 8 | 200 |
| NS8 | 8 | 8 | 200 |
| NS9 | 9 | 8 | 200 |
| NS10 | 10 | 8 | 200 |
| NS11 | 11 | 8 | 200 |
| NS12 | 12 | 8 | 200 |
| NS13 | 13 | 8 | 200 |

| | | | |
|---|---|---|---|
| NS14 | 14 | 8 | 200 |
| NS15 | 15 | 8 | 200 |
| NS16 | 16 | 8 | 200 |
| NS17 | 17 | 8 | 200 |
| NS18 | 18 | 8 | 200 |
| NS19 | 19 | 8 | 200 |
| NS20 | 20 | 8 | 200 |
| NS21 | 21 | 8 | 200 |

**Supplementary Table 3:** TWEC Training parameter for negative samples tuning

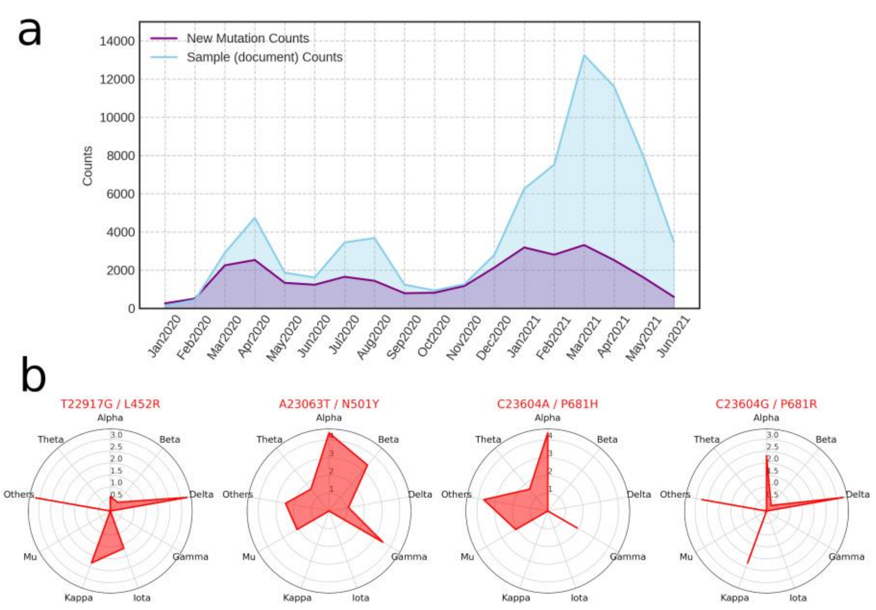


**Supplementary Fig 1: a)** Frequency of documents (i.e. genomic samples obtained from GISAID[8]) highlighted in blue, and frequency of new mutations highlighted in purple. **b)** Radial plot showing the distribution of four mutations of concern in samples classified according to the variant type. Inner contour lines correspond to frequency (scaled by log10).

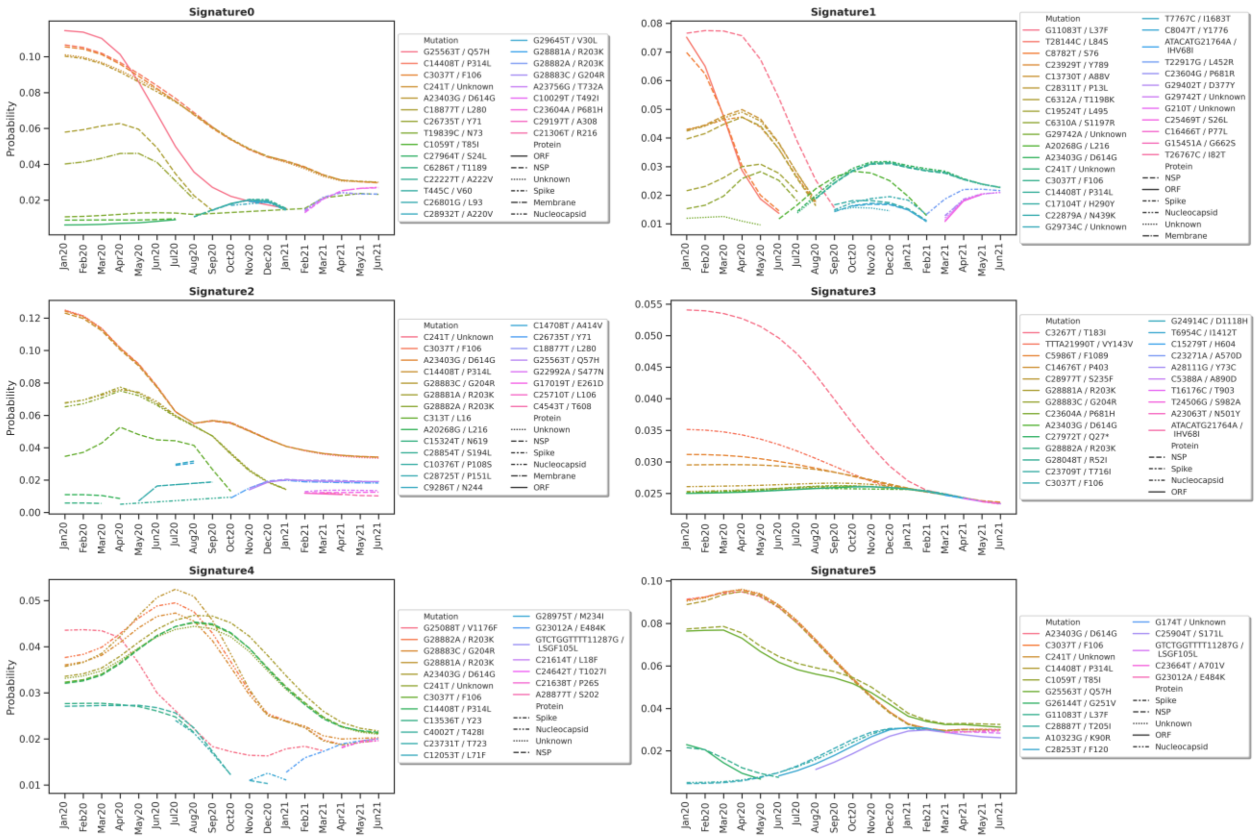


**Supplementary Fig2:** Diachronic shifts of word probabilities in each topic/signature

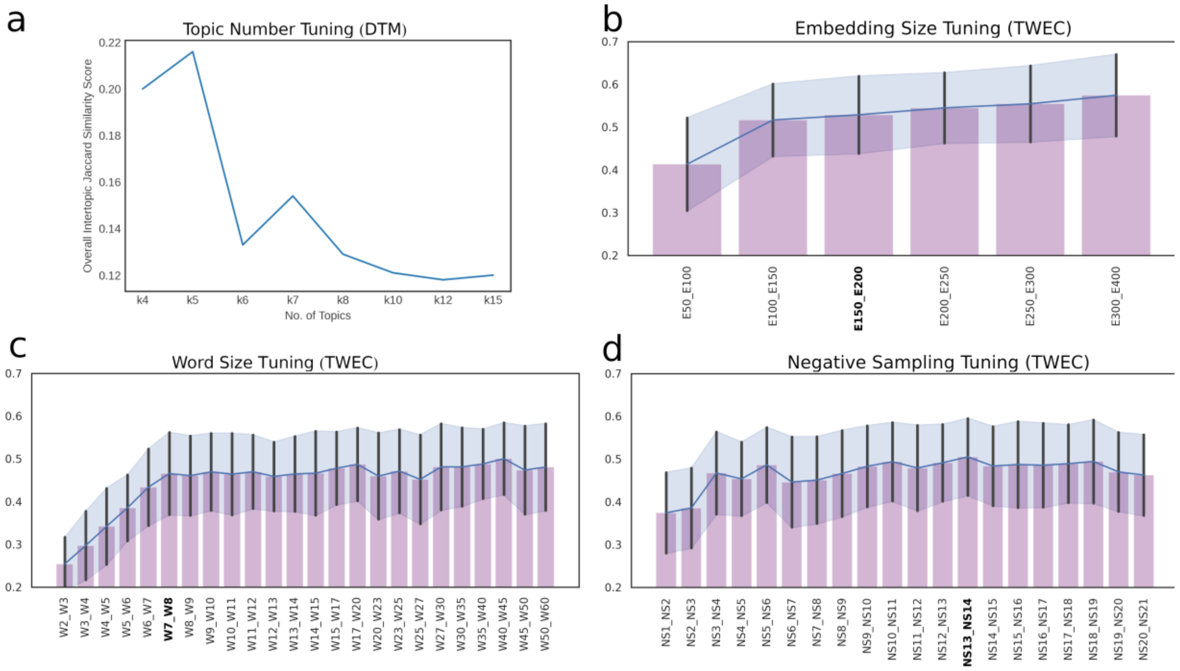

**Supplementary Fig3:** Hyperparameter tuning of DTM and TWEC models. **a)** Topic number selection via Jaccard (dis)similarity between topics of models trained on different topic numbers. For our study, we chose six as the topic number. Panels **b,c,d** correspond to hyperparameter optimisation for embedding size, word size and negative sampling (for Skipgram architecture of Word2Vec). The selected hyperparameter value is highlighted in bold on the X-axis of each panel.

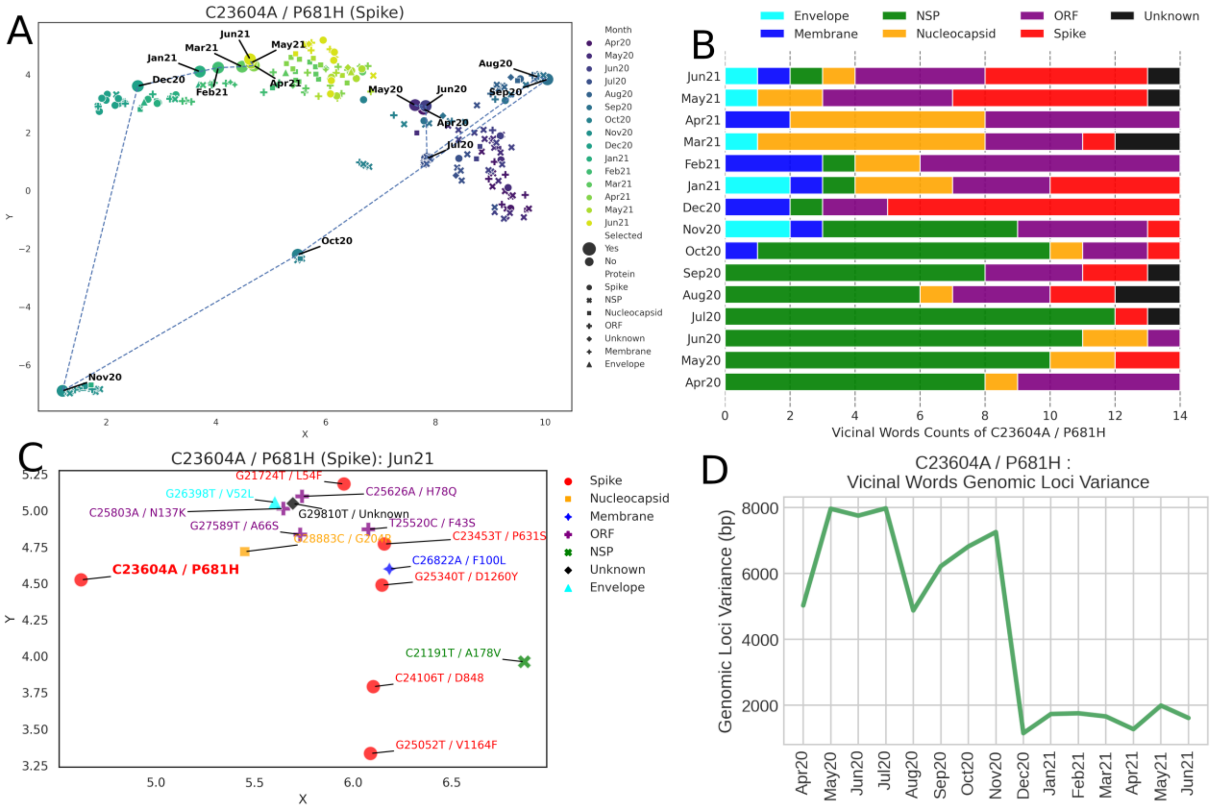


**Supplementary Fig4:** Semantic Drift of mutation C23604A / P681H (Spike) **a)** UMAP projection of word embedding of mutation C23604A and 15 most similar words for each time-slice from Apr2020 to Jun2021. The dotted line links the word embedding projection point for the mutation of concern. **b)** Neighbouring word distribution based on protein mutation for each time-slice. **c)** UMAP[9] projection of word embedding of mutation C23604A and 15 most similar words in the last time-slice. **d)** Standard deviations of genomic positions of neighbouring words of the mutation of concern.

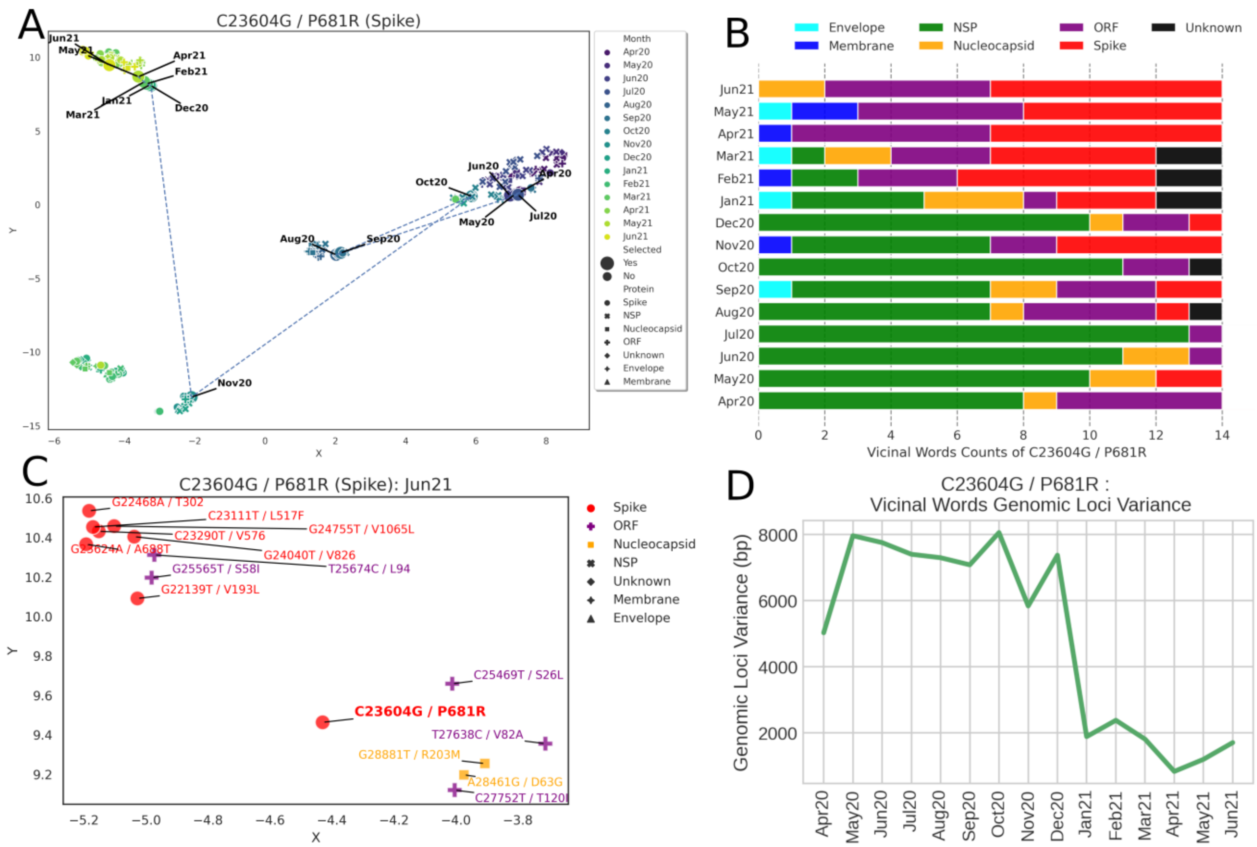

**Supplementary Fig5:** Semantic Drift of mutation C23604G / P681R (Spike) **a)** UMAP projection of word embedding of mutation C23604G and 15 most similar words for each time-slice from Apr2020 to Jun2021. The dotted line links the word embedding projection point for the mutation of concern. **b)** Neighbouring word distribution based on protein mutation for each time-slice. **c)** UMAP projection of word embedding of mutation C23604G and 15 most similar words in the last time-slice. **d)** Standard deviations of genomic positions of neighbouring words of the mutation of concern.

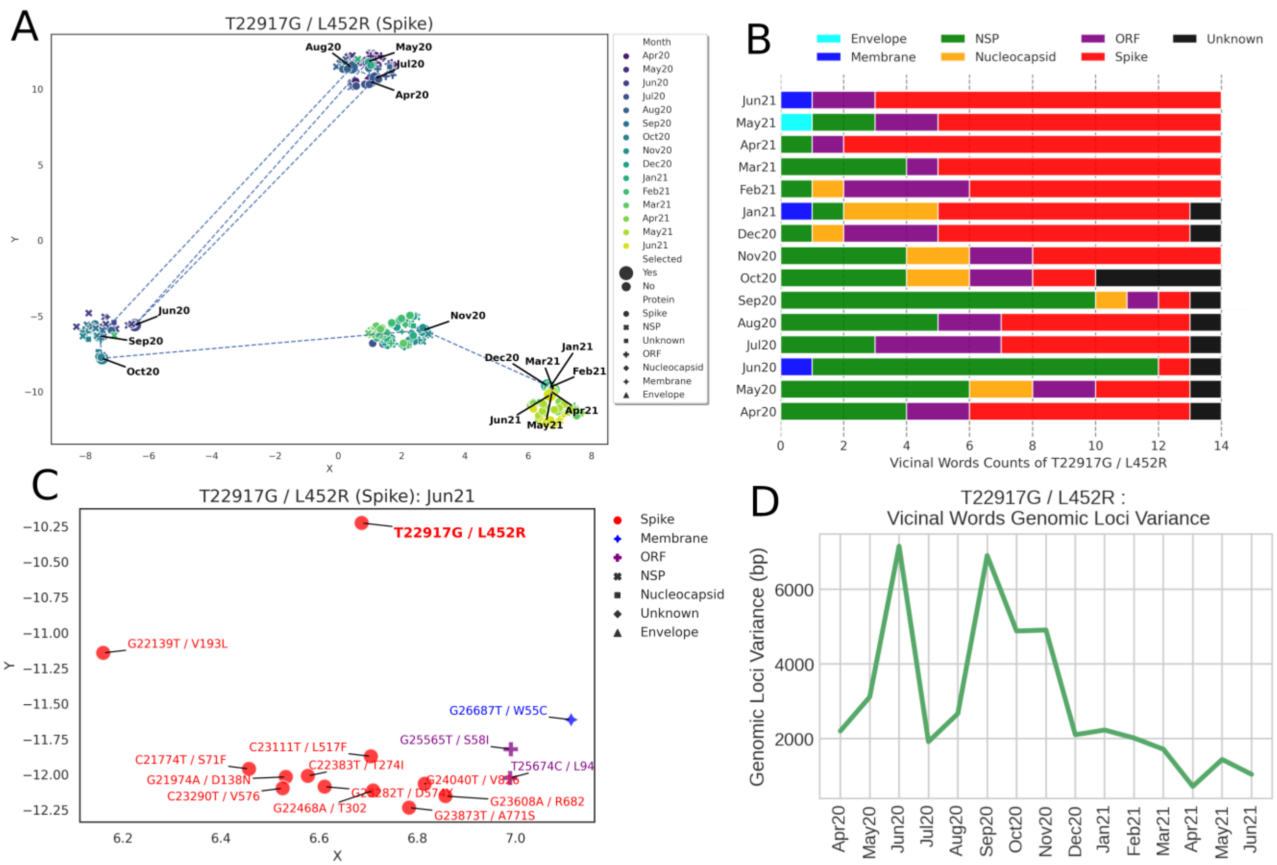


**Supplementary Fig6:** Semantic Drift of mutation T22917G / L452R (Spike) **a)** UMAP projection of word embedding of mutation T22917G and 15 most similar words for each time-slice from Apr2020 to Jun2021. The dotted line links the word embedding projection point for the mutation of concern. **b)** Neighbouring word distribution based on protein mutation for each time-slice. **c)** UMAP projection of word embedding of mutation T22917G and 15 most similar words in the last time-slice. **d)** Standard deviations of genomic positions of neighbouring words of the mutation of concern.

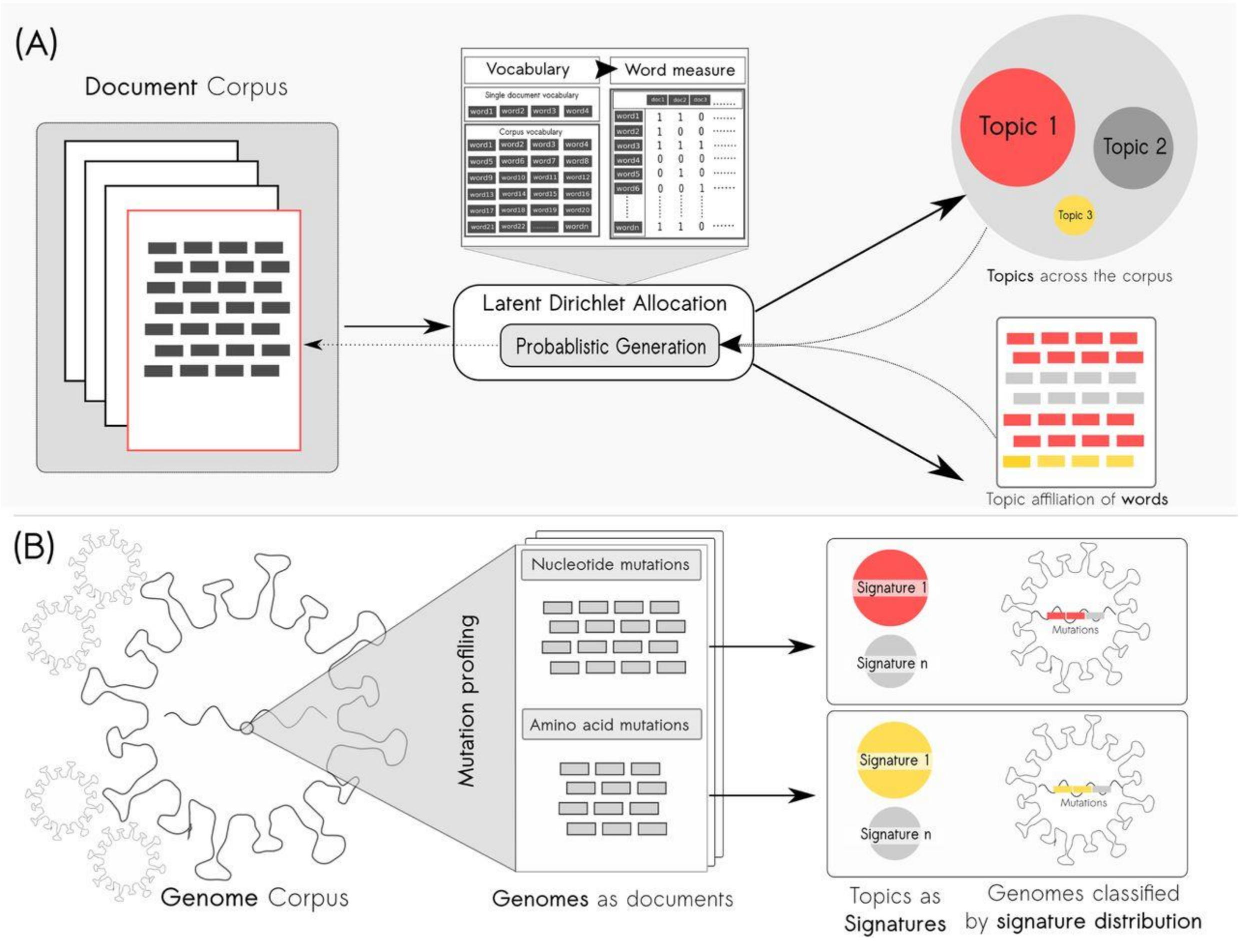


**Supplementary Fig7**: How to perceive SARS-CoV-2 genomes as documents[10]. **Panel A:** Classical approach towards topic modeling on large document corpus using the generative process of Latent Dirichlet Allocation (LDA). **Panel B:** Each SARS-CoV-2 genome with its mutation profile is treated as a document containing words in the form of their mutations with a potential to infer latent mutation signatures (topics).

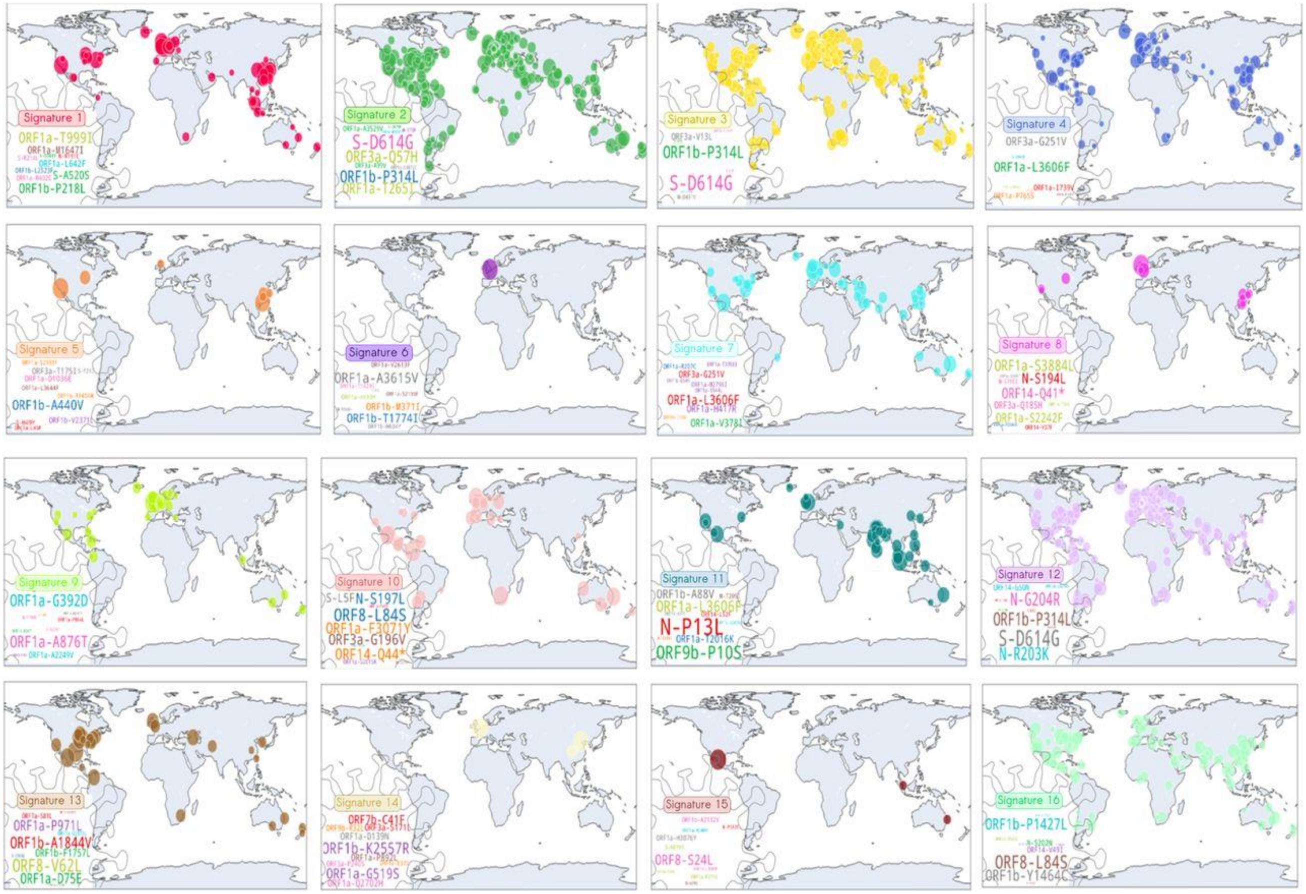


**Supplementary Fig8:** Geographical spread of putative signatures obtained previously from static LDA. Source[10]

1. Blei, D. M., Ng, A. Y. & Jordan, M. I. Latent Dirichlet Allocation. *J. Mach. Learn. Res.* **3**, 993–1022 (2003).

2. Liu, L., Tang, L., Dong, W., Yao, S. & Zhou, W. An overview of topic modeling and its current applications in bioinformatics. *SpringerPlus* **5**, 1608 (2016).

3. Zhang, Y. *et al.* Systematic identification of latent disease-gene associations from PubMed articles. *PLoS ONE* **13**, (2018).

4. Backenroth, D. *et al.* FUN-LDA: A Latent Dirichlet Allocation Model for Predicting Tissue-Specific Functional Effects of Noncoding Variation: Methods and Applications. *American Journal of Human Genetics* **102**, (2018).

5. Blei, D. M. & Lafferty, J. D. Dynamic topic models. in *ACM International Conference Proceeding Series* vol. 148 (2006).

6. Mikolov, T., Chen, K., Corrado, G. & Dean, J. Efficient estimation of word representations in vector space. in *1st International Conference on Learning Representations, ICLR 2013 - Workshop Track Proceedings* (2013).

7. di Carlo, V., Bianchi, F. & Palmonari, M. Training temporal word embeddings with a compass. in *33rd AAAI Conference on Artificial Intelligence, AAAI 2019, 31st Innovative Applications of Artificial Intelligence Conference, IAAI 2019 and the 9th AAAI Symposium on Educational Advances in Artificial Intelligence, EAAI 2019* (2019). doi:10.1609/aaai.v33i01.33016326.

8. Shu, Y. & McCauley, J. GISAID: Global initiative on sharing all influenza data – from vision to reality. *Eurosurveillance* vol. 22 (2017).

9. McInnes, L., Healy, J., Saul, N. & Großberger, L. UMAP: Uniform Manifold Approximation and Projection. *Journal of Open Source Software* **3**, (2018).

10. Nagpal, S., Srivastava, D. & Mande, S. S. What if we perceive SARS-CoV-2 genomes as documents? Topic modelling using Latent Dirichlet Allocation to identify mutation signatures and classify SARS-CoV-2 genomes. *bioRxiv* 2020.08.20.258772 (2020) doi:10.1101/2020.08.20.258772.
